# *FCGR3A* F158V Genotype Accelerates Progression to Chronic Alloimmune Injury in Antibody-Mediated Kidney Transplant Rejection: A Population-Based Cohort Study

**DOI:** 10.1101/2025.05.09.25327308

**Authors:** Thomas Vanhoutte, Karolien Wellekens, Priyanka Koshi, Jasper Callemeyn, Evert Cleenders, Angelica Paglizazzi, Elisabet Van Loon, Thibaut Vaulet, Brendan Keating, Alice Koenig, Olivier Thaunat, Georg Böhmig, Maarten Coemans, Dirk Kuypers, Maarten Naesens

## Abstract

**Background:** Evidence for involvement of NK cells and monocytes as prime agents in the process of human kidney transplant rejection is expanding quickly. These cells express FCGR3A, used for antibody-dependent cellular cytotoxicity (ADCC) after engagement with donor-specific antibodies on graft cells. A functional genetic polymorphism in the Fc fragment of IgG IIIa receptor (CD16a) gene (*FCGR3A*) results in a high-affinity (V158 allele) or low-affinity receptor (F158 allele).

**Methods:** We studied the impact of the *FCGR3A* F158V genotype on graft histology, function, and outcomes in a large, unselected observational cohort study of 1259 kidney transplants (40% F/F158, 46% F/V158 and 14% V/V158) with 5435 post-transplant transplant biopsies.

**Results:** Presence of at least one V158 allele was proportionally and significantly associated with a lower rate of C4d without rejection (F/V158, HR=0.51 [0.29-0.92], p=0.026; V/V158, HR=0.28 [0.08-0.90], p=0.033). Additionally, V/V158 was strongly associated with chronic active antibody-mediated rejection (HR=9.13 [1.89–44], p=0.006) and transplant glomerulopathy (HR=1.67 [1.01–2.75], p=0.046). The V/V158 genotype significantly associated with accelerated graft functional decline (−1.68 mL/min/y versus −0.74 mL/min/y (F/F158), p=0.034) and a higher rate of graft failure (HR=1.49 [1.01-2.20], p=0.045). This association was driven by transplants with post-transplant rejection.

**Conclusions:** Our findings suggest that FCGR3A affinity is key in AMR pathobiology. Low affinity of this receptor (F158 allele) might associate with abrogation of effective downstream ADCC, even in the presence of complement activation (C4d deposition), while high affinity of this receptor (V158 allele) might escalate AMR, accelerating evolution to more chronic AMR phenotypes (transplant glomerulopathy). This framework provides an immunobiological link between lower C4d without rejection rates, higher chronic active AMR/ transplant glomerulopathy rates and adverse graft outcomes associated with the *FCGR3A* V/V158 genotype.

**Key study findings:**

- V/V158 associates with higher rates of chronic active AMR/transplant glomerulopathy, and lower rates of C4d without rejection
- V/V158 drives faster graft decline and graft failure, especially in transplants complicated with rejection
- *FCGR3A* F158V might link complement activation and ADCC in AMR pathobiology, accelerating chronic alloimmune injury

## Introduction

Evidence on a pivotal role of CD16 (Fc fragment of IgG IIIa receptor, *FCGR3A*) expressing natural killer (NK) cells and monocytes as prime agents in kidney transplant rejection is expanding quickly, both with respect to their role in allorecognition and in secondary injury to allogeneic graft tissues. Biopsy-based transcriptomic analyses of bulk mRNA over the last decade have consistently shown enrichment of NK cell related transcripts in both donor-specific antibody (DSA) positive and DSA-negative microvascular inflammation (MVI), many of which are FCGR3A-inducible^1–4^. More recently, single cell RNA-sequencing data showed a significant correlation of only FCGR3A-positive NK cells and monocytes with intragraft inflammation severity^5^.

Recognition of DSA by NK cell and monocyte Fc gamma receptors (FCGRs) followed by antibody-dependent cellular cytotoxicity (ADCC) and graft injury constitutes an important pathway in kidney transplant rejection. NK cells mainly express the activating FCGR3A, while monocytes express activating FCGRs (FCGR1, FCGR2A and FCGR3A) as well as inhibitory FCGRs (FCGR2B)^6^. Within the hierarchy of, otherwise carefully balanced, receptors expressed on NK cells, FCGR3A takes center stage, as it lack inhibitory counterbalance and is the only receptor that can induce cytokine secretion without need of additional signaling^7^.

The *FCGR3A* gene displays a well-known nonsynonymous single nucleotide polymorphism (SNP) that substitutes phenylalanine for valine at position 158 (F158V) in the mature protein^8^. F158V enhances the affinity of FCGR3A for IgG1 and IgG3, in addition to gaining affinity for IgG2 and IgG4, and leads to higher NK cell surface expression and greater NK cell activation after antibody engagement^8–13^.

Several retrospective studies have reported on associations of F158V with incidence or severity of (antibody-mediated) rejection and graft failure (Supplementary Table 1)^10,11,14–19^. These studies vary in design (case-control; [often highly] selected; or unselected cohort), sample size (most being small) and granularity of clinical data (generally limited). While some of these studies associated the V158 allele with AMR-related histological changes, the associations with graft functional decline or failure were inconsistent.

We hypothesized that the affinity of FCGR3A, predicted by its F158V polymorphism, is key in the pathobiology, risk and severity of antibody-mediated rejection and outcome after kidney transplantation. To test this hypothesis, we studied the F158V genotype in relation to graft histology, functional decline and failure in a large, unselected and extensively phenotyped observational cohort study.

## Methods

### Study population and data collection

This unicentric observational cohort study included 1259 consecutive adult kidney transplants between March 2004 and September 2017 at the University Hospitals Leuven. Combined transplants or kidney transplants following a non-kidney organ transplant were excluded. Biopsies were performed at baseline, at specific protocol biopsy time points (3, 12, and 24 months post-transplantation) and when clinically indicated. Additionally, recipients transplanted before October 2005, November 2008, and January 2010 underwent protocol biopsies at 48, 36, and 60 months, respectively, as part of the center’s routine clinical protocol. Data collection ended on June 11, 2022. Prospectively collected clinical data was transferred from electronic health records to the research database. The clinical data from this observational cohort study (clinicaltrials.gov NCT06505200) was combined with newly generated genetic data (*FCGR3A* F158V genotype) on recipient DNA samples collected prospectively in the Biobank Renal Transplantation of the University Hospitals Leuven (clinicaltrials.gov NCT01331668). The Ethics committee of University Hospitals Leuven (S64006 and S64904) approved this project and all participants provided written informed consent. This research complied with the World Medical Association’s Declaration of Helsinki principles.

### Clinicopathological assessment of post-transplant biopsies

All biopsies performed between 2004 and 2017 in this cohort were prospectively catalogued in the electronic medical records, and retrospectively re-evaluated for Banff lesion scores. From 2017 onwards, Banff lesion scores were prospectively entered into the electronic medical records and transferred to the research database with the other prospectively collected clinical data. All adequate biopsies were reclassified to the Banff 2022 criteria, incorporating Banff lesion scores and information regarding HLA-DSA status. C4d staining in peritubular capillaries was performed using immunohistochemistry on frozen samples and was scored as follows: C4d0 (no staining), C4d1 (minimal staining, >0% but <25% of PTCs), C4d2 (focal staining, 25%-75% of PTCs), and C4d3 (diffuse staining, >75% of PTCs). In our center a C4d score of ≥2 has been validated as the threshold for C4d positivity^20^, with the threshold applicable to our staining method being concordant (89% accuracy) with the results of the evaluation by immunofluorescence on frozen samples.

### Detection of circulating (donor specific) HLA-antibodies

Screening for HLA-DSA was routinely performed before and after transplantation, coinciding with each biopsy, using a Luminex-based assay (HILA - Red Cross Flanders) to detect HLA-A, -B, -C, -DR, -DQ, and -DP antibodies with single antigen beads (SAB) tests (Immucor®). After our center transitioned from ELISA to the Luminex method in 2008, we re-analyzed prospectively biobanked sera (in the context of the Biobank Renal Transplantation NCT01331668) from specimens not initially evaluated with Luminex, as outlined recently^21^. The criteria for positivity were set at a median fluorescence intensity (MFI) exceeding 500 at the time of biopsy.

### FCGR3A genotyping

Genotyping of the *FCGR3A* gene for the F158V single nucleotide polymorphism (rs396991, 1:161544752 [GRCh38]) was performed using the Tx GWAS Array, designed by the iGENETRAiN consortium, based on Axiom Affymetrix technology, as described previously^22^.

### Statistical analysis

We report the study in concordance with the STROBE guidelines (Supplementary Table 2). R software (R version 4.4.2, “Puppy Cup”) was used for all statistical analyses. We employed a complete case analysis for handling missing data. Hypothesis testing was conducted two-tailed with a significance level set at p<0.05 for all analyses. Analyses were performed at the transplant level and stratified according to *FCGR3A* F158V genotype (F/F158, F/V158 or V/V158). Subcohorts were defined, starting from the first biopsy with specific rejection phenotypes: the “rejection cohort” (transplants with T-cell mediated rejection [TCMR]; active antibody-mediated rejection [AMR]; or microvascular inflammation [MVI], DSA-negative and C4d-negative) and “AMR/MVI cohort” (transplants with active AMR or MVI, DSA-negative and C4d-negative). Continuous variables were summarized as mean ± standard deviations (SD) or median ± interquartile range [IQR], and compared using ANOVA or Kruskal Wallis tests as appropriate. Categorical variables were summarized as frequencies and proportions (%) and compared using Chi-squared tests. Statistical results with 95% confidence intervals (CI) are reported with confidence bounds presented in square brackets.

#### Graft histology

Cox regression analysis was used to examine rates of Banff rejection phenotypes and transplant glomerulopathy. Analyses were performed using univariable and multivariable models, adjusted for recipient and donor age and sex, repeat transplantation, HLA mismatch and pretransplant HLA-DSA. Cumulative incidences were computed using the complement of the Kaplan-Meier method, with the log-rank test to assess differences. In a separate analysis, we studied the cumulative incidence of AMR phenotypes at the first qualifying biopsy within a defined AMR spectrum (C4d without evidence of rejection, probable AMR, full AMR [active AMR, chronic active AMR, and chronic AMR]), treating these phenotypes as competing risks using the Aalen-Johansen estimator and Gray’s test. Censoring was applied at the time of the last biopsy or administratively at 8 years after transplantation.

#### Graft function and outcome

Post-transplant evolution of graft function was studied using longitudinal linear mixed models with a fixed linear effect of post-transplantation time and CKD-EPI 2009 eGFR_cr_ as outcome. Random intercepts and linear random slopes were used to capture individual trajectories. All predictors were introduced as main effects, and tested for interaction with time. Given recovery of graft function during the first year after transplantation^23^, first year eGFR values were excluded from the analysis.

Graft failure rates and the risk of primary graft failure were studied using Cox regression and competing risks (death with functioning graft) (Fine and Gray) regression, respectively. The cumulative incidence of graft failure was computed using the Aalen-Johansen estimator, with Gray’s test to assess difference. Censoring was applied at the time of last follow-up, June 11 2022 or administratively at 12 years after transplantation.

Regression models within rejection subgroups were corrected for the time to first rejection. Analyses were performed using univariable and multivariable models, adjusted for recipient and donor age and sex, recipient BMI, repeat transplantation, donor type, cold ischemia time, HLA mismatch and pretransplant HLA-DSA.

In a separate analysis, the cumulative incidence and graft failure rate stratified by *FCGR3A* F158V genotype and starting from the first biopsy within the defined AMR spectrum (see above) was computed using the Aalen Johansen estimator and univariable Cox and competing risk regression, respectively.

## Results

### Characteristics of the study cohort

The study included 1259 individual transplants. The donor-recipient and post-transplant characteristics at time of transplantation are shown in Table 1. Recipients were mainly White Europeans (97%); donors were predominantly male (54%) and became donors after brain death (74%). Recipients almost invariably received modern era, post-transplant immunosuppression with tacrolimus (95%) and mycophenolate (96%). Median follow-up time after transplantation was 7.63 years [IQR: 4.86-11.24]. During follow-up, 215 grafts failed, and 358 recipients died. In total, 5526 biopsies were performed in 1249 transplants (498 F/F158, 573 F/V158 and 178 V/V158), of which 5435 were of adequate quality. *FCGR3A* genotype counts (%) were 502 (40%), 577 (46%) and 180 (14%) for F/F158, F/V158 and V/V158, respectively. This was in close agreement with frequencies expected under Hardy Weinberg equilibrium (Supplementary Table 3), and nearly identical to the frequencies reported previously^14^.

**Table 1.** Cohort characteristics: total and stratified per *FCGR3A* F158V genotype.

|  | *Total* | *F/F158* | *F/V158* | *V/V158* |
| --- | --- | --- | --- | --- |
| *Genotype count, N (%)* | 1259 (1.00) | 502 (0.40) | 577 (0.46) | 180 (0.14) |
| *Time of follow-up (days), median [IQR]* | 2787 [1836 - 4081.5] | 2822.5 [1775.5 - 4105.5] | 2734 [1866 - 4021] | 2998 [2011.25 - 4084.25] |
| *Cold ischemia time (h), mean ± SD* | 14.06 ± 4.79 | 14.27 ± 4.94 | 13.98 ± 4.64 | 13.71 ± 4.82 |
| *Repeat transplantation, N (%)* | 179 (0.14) | 67 (0.13) | 81 (0.14) | 31 (0.17) |
| *Delayed graft function, N (%)* | 213 (0.17) | 75 (0.15) | 105 (0.18) | 33 (0.18) |
| *Graft failure, N (%)* | 215 (0.17) | 74 (0.15) | 101 (0.18) | 40 (0.22) |
| *Death with functioning graft, N (%)* | 358 (0.28) | 138 (0.27) | 172 (0.3) | 48 (0.27) |
| *Recipient age (years), mean ± SD* | 54.05 ± 12.8 | 53.45 ± 12.95 | 54.89 ± 12.77 | 53.05 ± 12.36 |
| *Recipient BMI (kg/m<sup>2</sup>), mean ± SD* | 25.5 ± 4.5 | 25.54 ± 4.55 | 25.34 ± 4.32 | 25.91 ± 4.89 |
| *Male recipient, N (%)* | 795 (0.63) | 307 (0.61) | 372 (0.64) | 116 (0.64) |
| *African, N (%)* | 31 (0.02) | 19 (0.04) | 9 (0.02) | 3 (0.02) |
| *White Europeans, N (%)* | 1224 (0.97) | 482 (0.96) | 565 (0.98) | 177 (0.98) |
| *Hispanic, N (%)* | 1 (0) | 0 (0) | 1 (0) | 0 (0) |
| *Donor age (years), mean ± SD* | 48.5 ± 14.4 | 48.25 ± 14.08 | 49.4 ± 14.35 | 46.26 ± 15.25 |
| *Male donor, N (%)* | 676 (0.54) | 272 (0.54) | 304 (0.53) | 100 (0.56) |
| *Living donation, N (%)* | 89 (0.07) | 36 (0.07) | 43 (0.07) | 10 (0.06) |
| *Donation after cardiac death, N (%)* | 235 (0.19) | 102 (0.2) | 101 (0.18) | 32 (0.18) |
| *Donation after brain death, N (%)* | 935 (0.74) | 364 (0.73) | 433 (0.75) | 138 (0.77) |
| *HLA-A mismatch (broad), mean ± SD* | 0.83 ± 0.66 | 0.83 ± 0.66 | 0.84 ± 0.66 | 0.78 ± 0.64 |
| *HLA-B mismatch (broad), mean ± SD* | 0.97 ± 0.59 | 0.96 ± 0.57 | 0.99 ± 0.6 | 0.96 ± 0.57 |
| *HLA-DR mismatch (broad), mean ± SD* | 0.68 ± 0.55 | 0.66 ± 0.54 | 0.71 ± 0.55 | 0.62 ± 0.56 |
| *HLA-A/B/DR mismatch (broad), mean ± SD* | 2.47 ± 1.21 | 2.44 ± 1.18 | 2.54 ± 1.24 | 2.37 ± 1.19 |
| *Panel reactive antibody (%), mean ± SD* | 55.36 ± 32.17 | 52.34 ± 33.62 | 57.14 ± 31.58 | 57.39 ± 30.93 |
| *Overall pretransplant HLA-DSA, N (%)* | 100 (0.08) | 34 (0.07) | 53 (0.09) | 13 (0.07) |
| *Induction, N (%)* | 542 (0.43) | 223 (0.44) | 241 (0.42) | 78 (0.43) |
| *Basiliximab, N (%)* | 510 (0.41) | 211 (0.42) | 226 (0.39) | 73 (0.41) |
| *Tacrolimus, N (%)* | 1196 (0.95) | 474 (0.94) | 550 (0.95) | 172 (0.96) |
| *Mycophenolate mofetil, N (%)* | 1206 (0.96) | 481 (0.96) | 549 (0.95) | 176 (0.98) |

### *FCGR3A* F158V genotype, Banff rejection phenotypes and transplant glomerulopathy

Incidence rates of Banff rejection categories and transplant glomerulopathy per transplant and per biopsy according to *FCGR3A* genotype are shown in Table 2. Chronic active AMR occurred significantly more frequent in transplants with the V/V158 genotype as compared to F/V158 and F/F158 (3.9% vs. 0.9%, p=0.002, and 0.4% respectively, p=0.012). Conversely, C4d without rejection was significantly less frequent with increasing V158 alleles when compared to F/F158 (6% in F/F158 vs. 3.1% in F/V158 and 1.7% in V/V158, p=0.013).

**Table 2.** Incidence rates of Banff rejection phenotypes and transplant glomerulopathy per transplant and per biopsy: total and stratified per *FCGR3A* F158V genotype.

|  | *Total* | *F/F158* | *F/V158* | *V/V158* | *P value*<sup>¶</sup> |
| --- | --- | --- | --- | --- | --- |
| *Number of transplantations* | 1249 | 498 | 573 | 178 |  |
| *Borderline TCMR, N (%)* | 255 (0.204) | 100 (0.201) | 121 (0.211) | 34 (0.191) | 0,820 |
| *TCMR, N (%)* | 354 (0.283) | 143 (0.287) | 162 (0.283) | 49 (0.275) | 0,954 |
| *Chronic active TCMR, N (%)* | 19 (0.015) | 5 (0.01) | 9 (0.016) | 5 (0.028) | 0,238 |
| *DSA C4d- MVI, N (%)* | 169 (0.135) | 71 (0.143) | 75 (0.131) | 23 (0.129) | 0,828 |
| *C4d without rejection, N (%)* | 51 (0.041) | 30 (0.06) | 18 (0.031) | 3 (0.017) | **0,013** |
| *Probable AMR, N (%)* | 28 (0.022) | 6 (0.012) | 17 (0.03) | 5 (0.028) | 0,130 |
| *Active AMR, N (%)* | 97 (0.078) | 38 (0.076) | 43 (0.075) | 16 (0.09) | 0,803 |
| *Chronic active AMR, N (%)* | 14 (0.011) | 2 (0.004) | 5 (0.009) | 7 (0.039) | **<0,001** |
| *Chronic AMR, N (%)* | 15 (0.012) | 5 (0.01) | 7 (0.012) | 3 (0.017) | 0,772 |
| *No rejection, N (%)* | 1223 (0.979) | 493 (0.99) | 558 (0.974) | 172 (0.966) | 0,078 |
| *Transplant glomerulopathy, N (%)* | 129 (0.103) | 45 (0.09) | 60 (0.105) | 24 (0.135) | 0,244 |
| *Number of biopsies* | 5435 | 2199 | 2478 | 758 |  |
| *Borderline TCMR, N (%)* | 304 (0.056) | 117 (0.053) | 143 (0.058) | 44 (0.058) | 0,770 |
| *TCMR, N (%)* | 520 (0.096) | 206 (0.094) | 241 (0.097) | 73 (0.096) | 0,916 |
| *Chronic active TCMR, N (%)* | 22 (0.004) | 5 (0.002) | 11 (0.004) | 6 (0.008) | 0,099 |
| *DSA C4d- MVI, N (%)* | 259 (0.048) | 105 (0.048) | 116 (0.047) | 38 (0.05) | 0,932 |
| *C4d without rejection, N (%)* | 63 (0.012) | 36 (0.016) | 23 (0.009) | 4 (0.005) | **0,017** |
| *Probable AMR, N (%)* | 34 (0.006) | 8 (0.004) | 20 (0.008) | 6 (0.008) | 0,130 |
| *Active AMR, N (%)* | 158 (0.029) | 60 (0.027) | 77 (0.031) | 21 (0.028) | 0,722 |
| *Chronic active AMR, N (%)* | 17 (0.003) | 2 (0.001) | 7 (0.003) | 8 (0.011) | **<0,001** |
| *Chronic AMR, N (%)* | 17 (0.003) | 5 (0.002) | 7 (0.003) | 5 (0.007) | 0,173 |
| *No rejection, N (%)* | 4221 (0.777) | 1725 (0.784) | 1922 (0.776) | 574 (0.757) | 0,297 |
| *Transplant glomerulopathy, N (%)* | 167 (0.031) | 53 (0.024) | 80 (0.032) | 34 (0.045) | **0,017** |

<sup>¶</sup>P values from chi-squared tests comparing distributions across *FCGR3A* genotypes. Significant values are shown in **bold**.

Rates of Banff rejection categories according to *FCGR3A* F158V genotype (with F/F158 as reference category) were studied using Cox regression analysis (Figure 1, Supplementary Table S4). V/V158 associated with a higher rate of chronic active AMR (HR=9.13 [1.894-44.028], p=0.006). Additionally, presence of the V158 allele cumulatively associated with lower rates of C4d without rejection (F/V158, HR=0.51 [0.29-0.92], p=0,026; V/V158, HR=0.28 [0.08-0.90], p=0,033). Finally, V/V158 associated with rates of transplant glomerulopathy (cg=1-3), HR=1.67 [1.01–2.75] (p=0.046) and higher scores of transplant glomerulopathy (cg=2-3 vs. 0-1; HR=2.26 [1.03-4.98], p=0.043; cg = 3 vs. 0-2; HR=3.21 [1.16-8.84], p=0.024) (Supplementary Table S5). All associations remained significant in the multivariable models. Analyses of cumulative incidence functions for each of the Banff rejection categories and transplant glomerulopathy were consistent with the results of the Cox regression models (Supplementary Figure S1).

**Figure 1.**
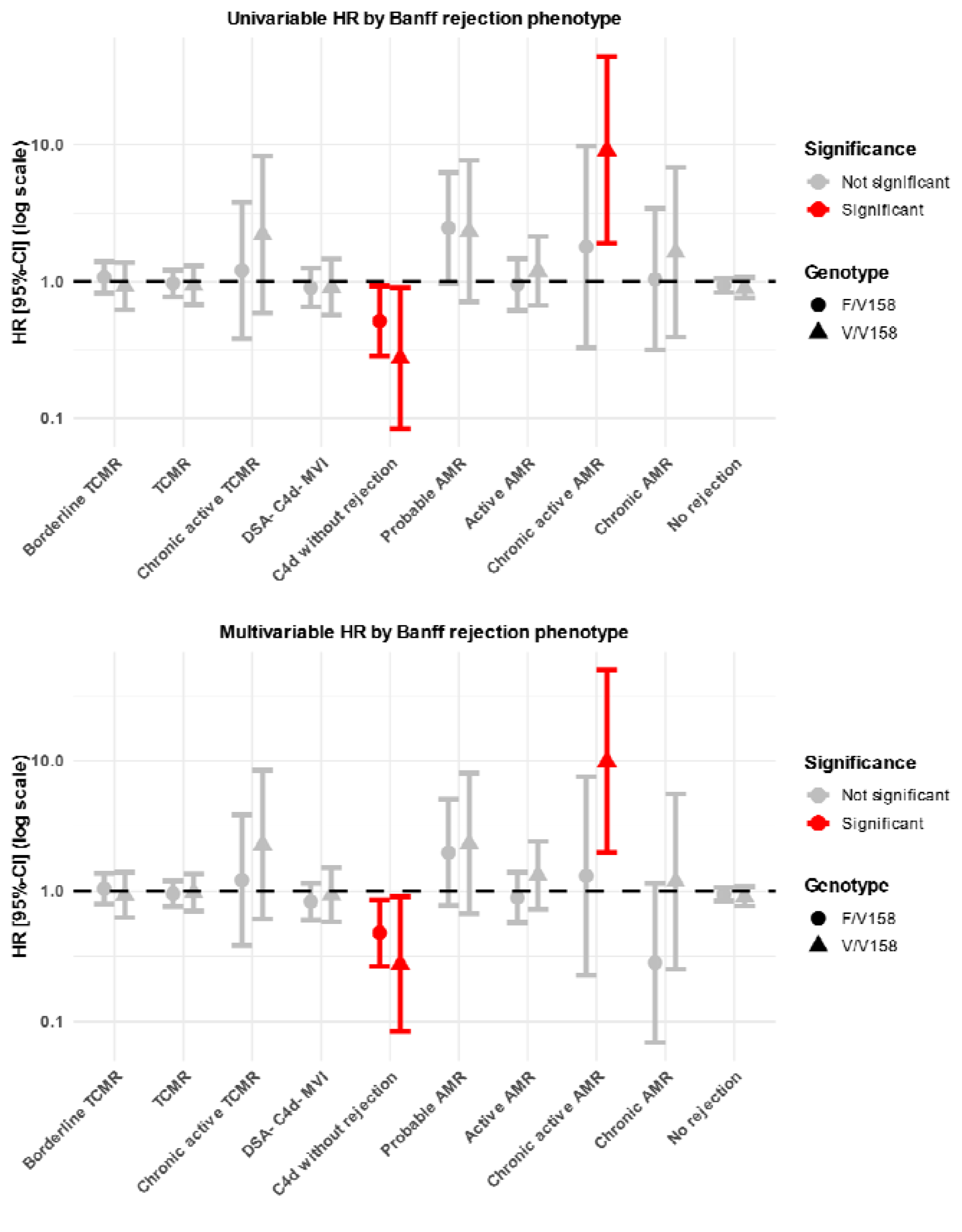
Results of univariable and multivariable Cox regression models examining the relation between *FCGR3A* F158V genotype and rates of Banff rejection phenotypes.

### *FCGR3A* F158V genotype and C4d without rejection in ABO compatible transplantation

In total, 51 transplants developed C4d without rejection: 30 (59%) in the F/F158 group, 18 (35%) in the F/V158 group and 3 (6%) in the V/V158 group. Of these, 47 were ABO-compatible, while 4 were ABO-incompatible living donor transplants that underwent pre-transplant desensitization. C4d deposition (without rejection) is observed in virtually all (early) biopsies in ABO-incompatible transplants because of persisting (mainly IgM^24^) ABO-antibodies, while it is a rare and ill-understood phenotype in ABO-compatible transplants. Assuming distinct pathophysiology, we excluded the 4 ABO-incompatible transplants from further analyses. After exclusion, the association between C4d without rejection and F/F158 remained significant (p=0.04), with a significantly higher cumulative incidence of C4d without rejection compared to F/V158 and V/V158 (log-rank test, p=0.034, Supplementary Figure S2).

Using the predefined competing risks framework in the Methods, we analyzed the cumulative incidence of C4d without rejection, probable AMR, and full AMR at the first biopsy qualifying for any of these phenotypes. This analysis was performed because C4d without rejection following probable AMR or full AMR phenotypes might indicate different underlying pathophysiological mechanisms such as AMR in (partial) remission. Cumulative incidence of C4d without rejection at first biopsy was 5.7% in F/F158, 2.7% in F/V158 and 1.4% in V/V158 (p=0.034); no differences in cumulative incidence of probable AMR (p=0.3) and full AMR (p=0.29) were found, except for chronic active AMR, which occurred significantly more frequent in V/V158 (4.2% in F/F158, 0.6% in F/V158 and 11.2% in V/V158, p=0.013), when this subgroup was extracted from full AMR.(Supplementary Table S6).

Next, we studied graft survival starting from the first biopsy with C4d without rejection, probable AMR or full AMR stratified by *FCGR3A* F158V genotype. It is tempting to hypothesize that this selection represents an enriched subcohort of transplants with indirect (histopathological C4d deposition) or direct (current or historic presence of circulating DSA leading to AMR-related histological changes) evidence of DSA interacting with graft endothelium. In this high-risk subcohort, the cumulative incidence of graft failure, as well as the rates of graft failure were significantly higher in the V/V158 group, compared to F/F158 group (52.9% [V/V158] vs. 22.5% [F/V158], p=0.032); HR=2.5 [1.15-5.44], p= 0.020; sHR=2.20 [1.02-4.76], p=0.046, respectively) (Supplementary Figure S3).

### *FCGR3A* F158V genotype independently associates with accelerated graft function decline, particularly in transplants with rejection

Results of univariable and multivariable linear mixed models examining the association of *FCGR3A* F158V with graft functional decline in the whole cohort, rejection cohort and AMR/MVI cohort are shown in Table 3 and Supplementary Tables S7 and S8. Predicted graft functional decline by FCGR3A genotype is shown in Figure 2.

**Table 3.**
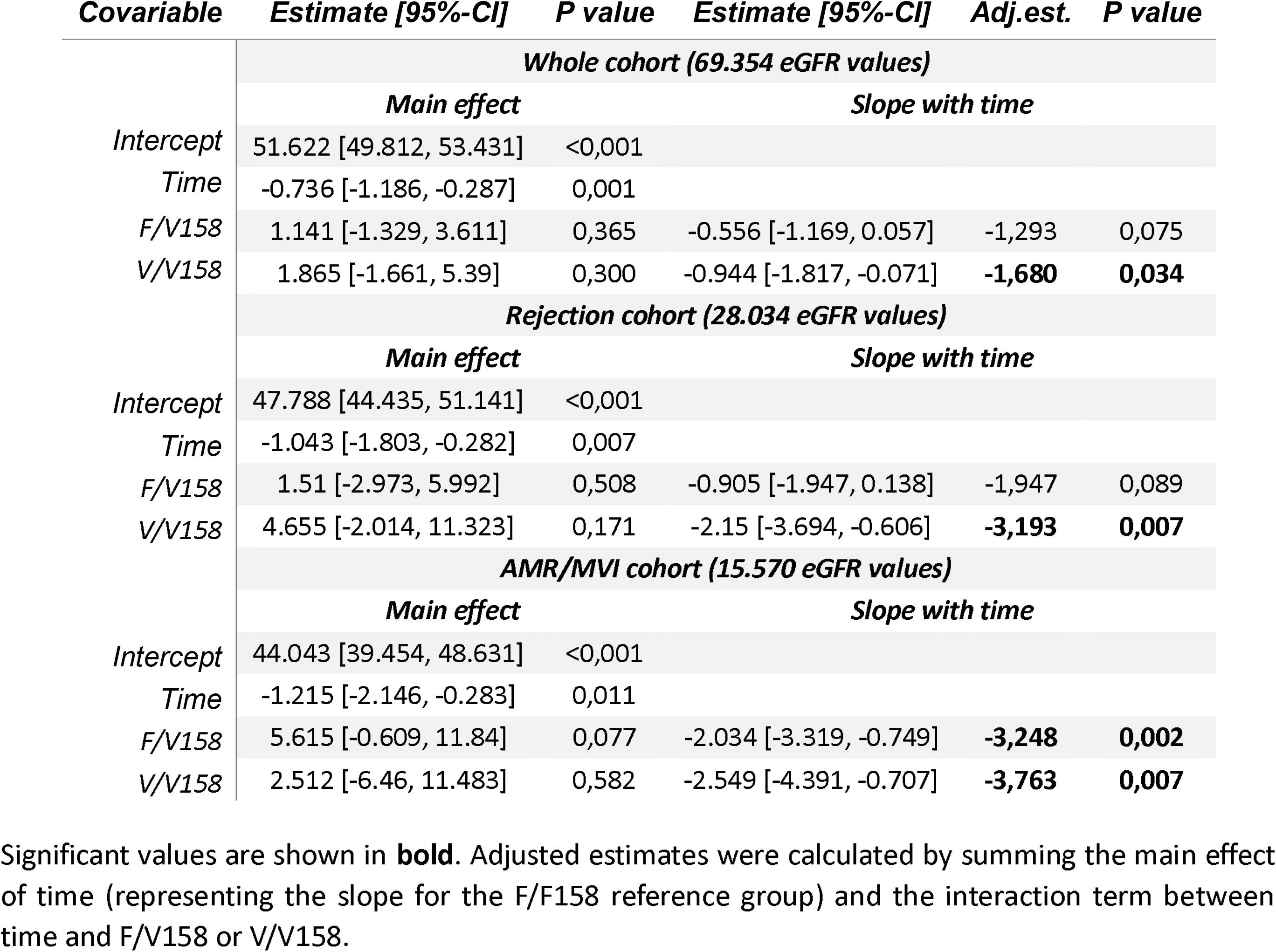
Results of univariable linear mixed models assessing the association of *FCGR3A* F158V genotype with graft functional decline in the whole cohort, rejection cohort and AMR/MVI cohort.

| *Covariable* | *Estimate [95%-CI]* | *P value* | *Estimate [95%-CI]* | *Adj.est.* | *P value* |
| --- | --- | --- | --- | --- | --- |
| **Whole cohort (69.354 eGFR values)** |  |  |  |  |  |
|  | **Main effect** |  | **Slope with time** |  |  |
| *Intercept* | 51.622 [49.812, 53.431] | <0,001 |  |  |  |
| *Time* | -0.736 [-1.186, -0.287] | 0,001 |  |  |  |
| *F/V158* | 1.141 [-1.329, 3.611] | 0,365 | -0.556 [-1.169, 0.057] | -1,293 | 0,075 |
| *V/V158* | 1.865 [-1.661, 5.39] | 0,300 | -0.944 [-1.817, -0.071] | **-1,680** | **0,034** |
| **Rejection cohort (28.034 eGFR values)** |  |  |  |  |  |
|  | **Main effect** |  | **Slope with time** |  |  |
| *Intercept* | 47.788 [44.435, 51.141] | <0,001 |  |  |  |
| *Time* | -1.043 [-1.803, -0.282] | 0,007 |  |  |  |
| *F/V158* | 1.51 [-2.973, 5.992] | 0,508 | -0.905 [-1.947, 0.138] | -1,947 | 0,089 |
| *V/V158* | 4.655 [-2.014, 11.323] | 0,171 | -2.15 [-3.694, -0.606] | **-3,193** | **0,007** |
| **AMR/MVI cohort (15.570 eGFR values)** |  |  |  |  |  |
|  | **Main effect** |  | **Slope with time** |  |  |
| *Intercept* | 44.043 [39.454, 48.631] | <0,001 |  |  |  |
| *Time* | -1.215 [-2.146, -0.283] | 0,011 |  |  |  |
| *F/V158* | 5.615 [-0.609, 11.84] | 0,077 | -2.034 [-3.319, -0.749] | **-3,248** | **0,002** |
| *V/V158* | 2.512 [-6.46, 11.483] | 0,582 | -2.549 [-4.391, -0.707] | **-3,763** | **0,007** |

Significant values are shown in **bold**. Adjusted estimates were calculated by summing the main effect of time (representing the slope for the F/F158 reference group) and the interaction term between time and F/V158 or V/V158.

**Figure 2.**
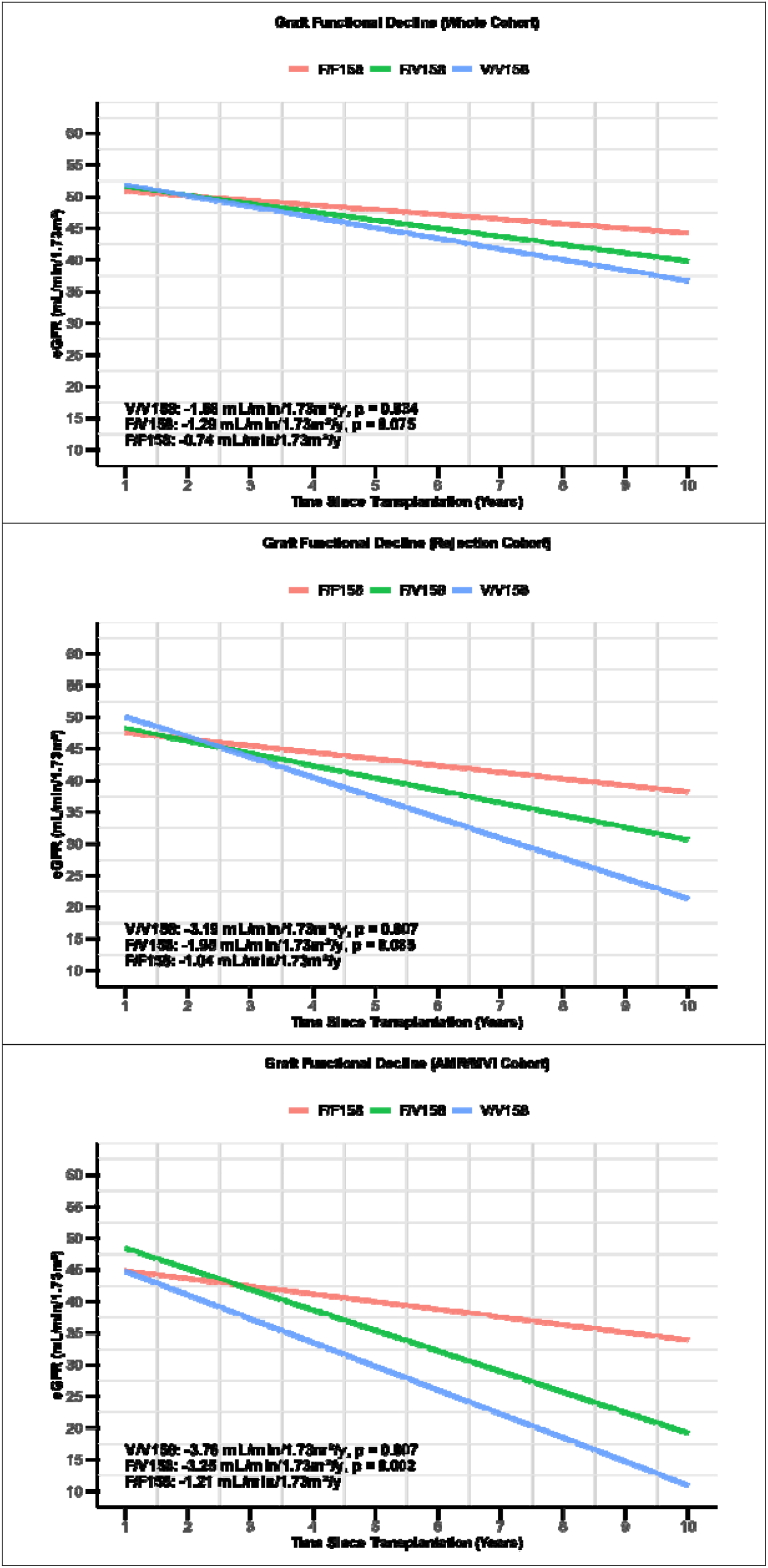
Predicted graft functional decline stratified by *FCGR3A* F158V genotype based on univariable linear regression. Predicted graft functional decline stratified by *FCGR3A* F158V genotype based on univariable linear regression. The top panel shows the whole cohort, the middle panel represents the rejection cohort, and the bottom panel depicts the AMR/MVI cohort. Lines indicate predicted estimated glomerular filtration rate (eGFR) over time (years post-transplant) for each genotype: F/F158 in red (reference), F/V158 in green, and V/V158 in blue. Slopes represent the rate of eGFR decline per year, with p-values shown for differences compared to the F/F158 reference group.

In univariable analysis, V/V158 significantly associated with a faster eGFR decline of −1.68 mL/min/1.73m^2^/year (p=0.034), while F/V158 associated with a borderline faster eGFR decline of −1.29 mL/min/1.73m^2^/year (p=0.075) vs. −0.74 mL/min/1.73m^2^/year associated with F/F158. Acceleration of graft functional decline associated with V/V158 vs. F/F158 became more pronounced in univariable linear mixed models based on data from the rejection (−3.19 mL/min/1.73m^2^/year vs. −1.04 mL/min/1.73m^2^/year; p=0.007) and AMR/MVI cohorts (−3.76 mL/min/1.73m^2^/year vs. 1.22 mL/min/1.73m^2^/year; p=0.007), corresponding to a 2.28 times, 3.06 and 3.1 times faster decline in the whole cohort, rejection cohort and AMR/MVI cohort, respectively. All these associations remained significant in the multivariable models. Interestingly, a dose-dependent effect of increasing V158 allele within the F/V158 genotype with faster eGFR decline was found versus the F/F158 genotype with steeper graft functional decline for every V158 allele added.

### *FCGR3A* F158V genotype independently associates with graft failure, particularly in transplants with rejection

The cumulative incidence of graft failure stratified by *FCG3RA* F158V genotype in the whole cohort, rejection cohort and AMR/MVI cohort is shown in Figure 3. Estimates for graft failure at 12 years for F/F158, F/V158 and V/V158 were 17.5%, 19.5% and 26.5% in the whole cohort (F/F158 vs. V/V158, p=0.04); 21.7%, 28.9% and 41.1% in the rejection cohort (F/F158 vs V/V158, p=0.01) and 23.4%, 33.5% and 46.1% in the AMR/MVI cohort (F/F158 vs V/V,158, p=0.03). Compared to F/F158, V/V158 had an estimated 1.5-fold, 1.9-fold, and 2.0-fold higher cumulative incidence of graft failure at 12 years, respectively. Notably, across all cohorts, estimates for graft failure increased in a dose-dependent manner with the number of V158 alleles.

**Figure 3.**
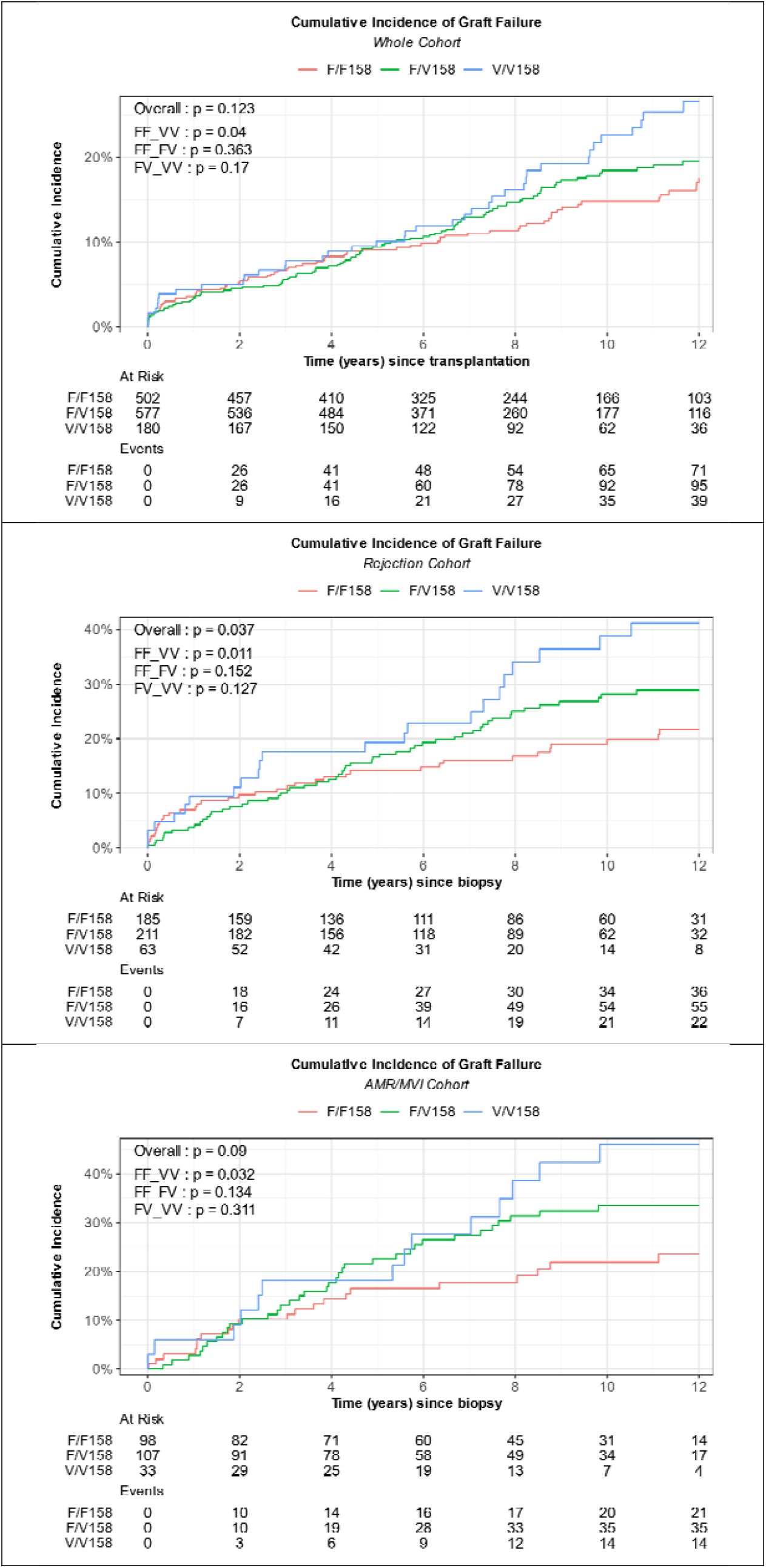
Cumulative incidence of graft failure in the whole cohort, rejection cohort and AMR/MVI cohort stratified by *FCGR3A* F158V genotype. Cumulative incidence of graft failure estimated using the Aalen-Johansen method, stratified by *FCGR3A* F158V genotype in the full cohort, rejection cohort, and AMR/MVI cohort. P-values are shown for Gray’s test (overall) and pairwise comparisons.

Results for univariable Cox regression analysis are presented in Table 4. V/V158 significantly associated with higher graft failure rates compared to F/F158. This association was primarily driven by transplants with rejection (whole cohort, HR=1.49 [1.01-2.20], p=0,045; rejection cohort, HR=2.00 [1.18-3.41], p=0,010; AMR/MVI cohort, HR=2.02 [1.03-3.98], p=0,041). In multivariable Cox regression analysis, the V/V158 genotype remained significantly and independently associated with higher graft failure rates in the whole cohort (HR=1.55 [1.05-2.29], p=0,029), rejection cohort (HR=1.88 [1.1-3.21], p=0.021) and AMR/MVI cohort (HR=1.97 [0.99-3.91], p=0.054) (Supplementary Table 9). Furthermore, in the rejection and AMR/MVI cohorts the V/V158 genotype showed the strongest association with graft failure among the tested covariables. Competing risk models yielded consistent, statistically significant results with similar effect sizes (Table 4 and Supplementary Table S9).

**Table 4.** Results of univariable Cox and competing risk regression models assessing the association between FCGR3A F158V and various transplant-relevant covariables with graft failure.

|  | Covariable | Whole cohort<br>TX=1259, events=205 |  | Rejection cohort<br>TX=459, events=113 |  | AMR/MVI cohort<br>TX=239, events=70 |  |
| --- | --- | --- | --- | --- | --- | --- | --- |
|  |  | HR [95%-CI] | P value | HR [95%-CI] | P value | HR [95%-CI] | P value |
| Cox regression | Repeat TX | 1.549 [1.097 - 2.189] | **0,013** | 1.812 [1.191 - 2.759] | **0,006** | 1.984 [1.217 - 3.235] | **0,006** |
|  | Recipient age/10y | 0.946 [0.850 - 1.053] | 0,310 | 0.837 [0.734 - 0.955] | **0,008** | 0.771 [0.657 - 0.904] | **0,001** |
|  | Male recipient | 0.747 [0.567 - 0.984] | **0,038** | 0.839 [0.580 - 1.215] | 0,354 | 0.628 [0.392 - 1.006] | **0,053** |
|  | Recipient BMI/kg/m<sup>2</sup> | 0.989 [0.958 - 1.021] | 0,479 | 1.001 [0.960 - 1.043] | 0,970 | 0.990 [0.938 - 1.045] | 0,712 |
|  | CIT/5h | 1.108 [0.984 - 1.247] | 0,090 | 1.004 [0.853 - 1.181] | 0,962 | 1.015 [0.843 - 1.223] | 0,874 |
|  | Donor age/10y | 1.152 [1.040 - 1.277] | **0,007** | 1.037 [0.905 - 1.188] | 0,605 | 0.840 [0.716 - 0.986] | **0,033** |
|  | Male donor | 1.022 [0.776 - 1.345] | 0,877 | 0.987 [0.680 - 1.432] | 0,944 | 1.334 [0.829 - 2.147] | 0,235 |
|  | Pretransplant HLA-DSA | 2.563 [1.765 - 3.721] | **<0.001** | 2.068 [1.336 - 3.202] | **0,001** | 2.105 [1.287 - 3.444] | **0,003** |
|  | HLA mismatch | 1.098 [0.983 - 1.228] | 0,098 | 1.094 [0.940 - 1.273] | 0,244 | 0.878 [0.725 - 1.062] | 0,179 |
|  | F/V158 | 1.174 [0.863 - 1.596] | 0,307 | 1.346 [0.883 - 2.051] | 0,167 | 1.515 [0.881 - 2.608] | 0,133 |
|  | V/V158 | 1.491 [1.009 - 2.203] | **0,045** | 2.003 [1.177 - 3.407] | **0,010** | 2.023 [1.028 - 3.979] | **0,041** |
| Competing risk regression | Repeat TX | 1.541 [1.089 - 2.183] | **0,015** | 1.883 [1.248 - 2.843] | **0,003** | 2.098 [1.295 - 3.397] | **0,003** |
|  | Recipient age/10y | 0.877 [0.786 - 0.978] | **0,019** | 0.779 [0.680 - 0.893] | **0,000** | 0.727 [0.621 - 0.852] | **0,000** |
|  | Male recipient | 0.733 [0.556 - 0.965] | **0,027** | 0.807 [0.556 - 1.170] | 0,260 | 0.626 [0.391 - 1.001] | **0,051** |
|  | Recipient BMI/kg/m<sup>2</sup> | 0.984 [0.950 - 1.020] | 0,380 | 0.999 [0.956 - 1.043] | 0,950 | 0.987 [0.929 - 1.050] | 0,690 |
|  | CIT/5h | 1.115 [0.995 - 1.250] | 0,062 | 1.010 [0.865 - 1.178] | 0,900 | 1.004 [0.852 - 1.185] | 0,960 |
|  | Donor age/10y | 1.111 [0.994 - 1.242] | 0,063 | 0.993 [0.858 - 1.149] | 0,920 | 0.818 [0.697 - 0.959] | **0,014** |
|  | Male donor | 1.021 [0.776 - 1.343] | 0,880 | 1.028 [0.709 - 1.492] | 0,880 | 1.389 [0.866 - 2.227] | 0,170 |
|  | Pretransplant HLA-DSA | 2.511 [1.726 - 3.652] | **<0.001** | 2.027 [1.318 - 3.116] | **0,001** | 2.111 [1.303 - 3.418] | **0,002** |
|  | HLA mismatch | 1.069 [0.944 - 1.210] | 0,290 | 1.078 [0.921 - 1.260] | 0,350 | 0.875 [0.723 - 1.060] | 0,170 |
|  | F/V158 | 1.155 [0.849 - 1.572] | 0,360 | 1.350 [0.885 - 2.060] | 0,160 | 1.518 [0.885 - 2.603] | 0,130 |
|  | V/V158 | 1.510 [1.026 - 2.222] | **0,036** | 1.941 [1.141 - 3.304] | **0,014** | 2.112 [1.092 - 4.083] | **0,026** |

Univariable Cox and competing risk regression analysis for the rejection and AMR/MVI cohort was corrected for time to first rejection. Significant values are shown in **bold**.

## Discussion

In this large, observational cohort study, *FCGR3A* F158V showed significant and independent associations with graft rejection phenotypes, functional decline, and outcome, confirming its nature as a significant risk genotype.

An interesting finding in our study is the significant association of V158 with higher rates of chronic active AMR and transplant glomerulopathy and lower rates of C4d without rejection. In ABO-incompatible transplants, >25% C4d positivity in the peritubular capillaries occurs in up to 94% of biopsies, and is considered a state of accommodation associated with excellent graft outcomes^25^. Conversely, in ABO-compatible grafts, this phenotype is rare (representing 2% of all biopsies performed) and more enigmatic^26^. Prior studies suggest that it is not associated with rapid graft functional decline, and only few patients progress to AMR within one year, anti-rejection treatment could improve kidney function, and some patients develop AMR at a later point in time, suggesting that at least in some, it might represent a state of “smoldering” AMR^26,27^.

C4d is considered the histological substrate for the presence of DSA, as DSA bound to donor-antigens activate complement with subsequent deposition of C4d that becomes covalently attached to donor-epithelium with a strong thioester bond, even long after the DSA have disappeared^28^. Mechanistically, dissociation between C4d deposition and rejection could be explained by partial activation of the tightly regulated complement system (with the cascade progressing no further than C3 convertase), elevated expression of complement-regulatory proteins on the graft endothelium or activation through the lectin pathway, which does not constitute a true allo-immune response^26,29^.

Our study suggests that C4d without rejection may also reflect reduced engagement of DSA-donor antigen immune complexes by recipient NK cells/monocytes expressing the low-affinity F158 allele, resulting only in complement activation and C4d deposition, without effective downstream ADCC (Supplemental figure 4). Conversely, recipient NK cells/monocytes expressing the high-affinity V158 allele are expected to zealously target these immune complexes, become activated, secrete inflammatory cytokines and exhibit ADCC towards the graft endothelium, more quickly escalating in mature/chronic (active) AMR rejection phenotypes and transplant glomerulopathy. Theoretically, while grafts cells could mitigate complement-mediated (alloimmune) injury with mechanisms present under normal physiological conditions (e.g., serum and surface-bound complement-regulatory proteins), absence of such normal biological defensive mechanisms against ADCC might render this a predominant mechanism of graft injury in AMR. This also resonates with studies showing that despite prevention of early acute AMR, chronic AMR can still occur in patients treated with complement blockade^30,31^.

Our study is one of the first to examine the evolution of graft functional decline according to the *FCGR3A* F158V genotype^19^. One previous study did not find a relation between *FCGR3A* F158V genotype and graft functional decline, possibly resulting from a smaller sample size and/or shorter follow-up^19^. Importantly, associations with post-transplant eGFR slopes provides a firmer biological basis for a *bona fide* effect when compared with the dichotomous outcome of graft failure. We found a significantly steeper slope of graft functional decline associated with the V/V158 genotype, independent from the effect of other transplant-relevant covariables. This association was driven by transplant with rejection, as the slopes became steeper when limiting the analysis to data from the rejection and AMR/MVI cohorts.

Finally, we found that V/V158 significantly and independently associated with higher graft failure rates. This association was again driven by transplant with rejection, reflected by increasing HRs in the rejection and AMR/MVI cohorts. Interestingly, the strongest association between the V/V158 genotype and graft survival was observed in our ‘high-risk’ cohort—transplants with direct or indirect evidence of DSA—suggesting that the *FCGR3A* F158V effect is linked to the presence of DSA. The impact of *FCGR3A* F158V genotype on graft failure emerged only after long-term follow-up, with survival curves diverging beyond five years post-transplantation. This may explain inconsistent findings in the literature, as studies with shorter follow-up failed to detect and effect (e.g. Diebold *et al*^*19*^), while longer-term studies observed decreased graft survival (e.g. Bailly *et al*^*18*^). Importantly, our study also indicates that the immunological risk associated with the high-functional V158 allele is concentrated in V/V158 homozygotes. Because of power issues, in many studies the F158 or V158 alleles were lumped together for comparative analysis, obscuring this finding.

Despite a large and extensively clinically phenotyped cohort with long-term follow-up, our study has several limitations. First, this is a single-center study, and external validation of these findings in multi-center studies is needed. Second, given the retrospective study design, there is the potential residual risk of bias and confounding of study findings, notwithstanding rigorous and careful application of statistical analyses. Third, 97% of the recipients in our cohort were White Europeans, and in addition to *FCGR3A* genotype distributions, graft outcomes or histological associations might not be generalizable to populations of different genetic ancestry. Fourth, as event rates for rare Banff rejection categories, and graft outcomes, especially in subcohorts were low, the possibility of chance associations remain. Fifth, histological associations were based on conventional histomorphology only, lacking the high-resolution power of biopsy-based transcriptomics, which were not available for this study.

In conclusion, our study demonstrates that the *FCGR3A* F158V polymorphism is a significant and independent risk factor for adverse outcomes in a large, observational cohort. We suggest that the affinity of NK cell/monocyte expressed FCGR3A predicted by the F158V polymorphism is key in AMR pathobiology: low affinity of this receptor (F158) might dissociate complement activation (hallmarked byC4d deposition) from effective downstream ADCC, while high affinity of this receptor (V158) might lead to accelerated escalation of AMR to chronic (active) phenotypes (hallmarked by transplant glomerulopathy). This mechanism provides an immunobiological link between lower rates of C4d without rejection, higher rates of chronic active AMR/transplant glomerulopathy and worse graft outcomes associated with the *FCGR3A* V/V158 genotype.

## Data Availability

The data supporting the findings of this study can be obtained upon reasonable request from the corresponding author, Maarten Naesens. However, these data cannot be made publicly available due to certain restrictions, such as containing sensitive information that could potentially compromise the privacy of research participants.

## Acknowledgements

We express our gratitude to the clinicians, surgeons, nursing staff, and patients of the University Hospitals Leuven, the lab technicians of the Nephrology and Renal Transplantation Research Group (KU Leuven) as well as the centers collaborating in the Leuven Collaborative Group for Nephrology (LSGN).

## Author contributions

TVH and MN designed the study. TVH, KW, PK, EV, JC, DK, and MN were involved in data collection and curation. TVH performed all analyses with input from MN. TVH and MN wrote the manuscript. All authors critically reviewed the manuscript and provided important intellectual input.

## Data sharing statement

The data supporting the findings of this study can be obtained upon request from the corresponding author, MN. However, these data cannot be made publicly available due to certain restrictions, such as containing sensitive information that could potentially compromise the privacy of research participants.

## Supplementary appendix

Supplementary tables and figures are provided in a separate PDF file. These include additional data, statistical analyses, and detailed results that support the findings of the manuscript.

