## Supplementary Appendix for "*FCGR3A* F158V Genotype Accelerates Progression to Chronic Alloimmune Injury in Antibody-Mediated Kidney Transplant Rejection: A Population-Based Cohort Study"

**SUPPLEMENTARY MATERIALS**

*Supplementary Tables*

**Table S1. Summary of retrospective studies reporting on associations between FCGR3A F158V and histology or graft outcomes.**

**Table S2. STROBE Statement.**

**Table S3. Hardy Weinberg equilibrium.**

**Table S4. Results of univariable and multivariable Cox regression analysis assessing the rates of Banff rejection categories according to *FCGR3A* F158V and transplant-relevant covariables.**

**Table S5. Results of univariable and multivariable Cox regression analysis assessing the rates of transplant glomerulopathy (cg) according to *FCGR3A* F158V and transplant-relevant covariables at different grades of cg (presence of lesion [cg=1-3 versus 0], medium grade lesion [cg=2,3 versus 0, 1 or high-grade lesion [cg=3 versus 0, 1, 2]).**

**Table S6. Cumulative incidence of C4d without rejection, probable AMR and full AMR (active AMR, chronic active AMR and chronic AMR) at first biopsy considering these phenotypes as competing risks.**

**Table S7. Results of univariable linear mixed models assessing the association of graft functional decline with *FCGR3A* F158V genotype and transplant-relevant covariables in the whole cohort, rejection cohort and AMR/MVI cohort.**

**Table S8. Results of multivariable linear mixed models assessing the association of graft functional decline with *FCGR3A* F158V genotype and transplant-relevant covariables in the whole cohort, rejection cohort and AMR/MVI cohort.**

**Table S9. Results of multivariable Cox and competing risk regression models assessing the association between *FCGR3A* F158V genotype and transplant-relevant covariables with graft failure in the whole cohort, rejection cohort and AMR/MVI cohort.**

*Supplementary Figures*

**Figure S1. Cumulative incidence of Banff rejection categories stratified by *FCGR3A* F158V genotype**

**Figure S2. Cumulative incidence of C4d without rejection stratified by *FCGR3A* F158V genotype with (upper panel) and without (lower panel) ABO-incompatible grafts.**

**Figure S3. Cumulative incidence of graft failure in the AMR spectrum cohort (C4d without rejection, probable AMR, active AMR, chronic active AMR and chronic AMR) stratified by *FCGR3A* F158V genotype.**

**Figure S4. Conceptual figure depicting dissociation/association of complement activation and effective ADCC according to *FCGR3A* F158V genotype in antibody-mediated kidney transplant rejection.**

**Table S1. Summary of retrospective studies reporting on associations between FCGR3A F158V and histology or graft outcomes.**

| Study reference | Sample size/ study design | Histology | Graft outcome |
|---|---|---|---|
| Arnold ML, et al. Am J Transplant. 2018. PMID: 29478298. | 85 DSA+ KTRs/ selected cohort | V158 allele associated with higher rate (and extent) of ptc (ptc score ≥1: 53.6% vs 25.9%; P = .018) | No effect on graft survival or graft function decline |
| Litjens N, et al. Sci Rep. 2021. PMID: 33846428. | 133 KTRs with chronic active AMR, 116 controls/ case control | V/V158 genotype associated with a higher g score | V/V158 genotype was an independent risk factor (HR 1.98; P = 0.04) for decreased graft survival |
| Wahrmann M, et al. Front Immunol. 2021. PMID: 34497614. | 1,940 KTRs/ unselected cohort; 799 F/F158 (0.41), 884 F/V158 (0.46), 257 V/V158 (0.13) | NA | No effect on graft survival |
| Vietzen H, et al. Front Immunol. 2022. PMID: 35401541 | 86 DSA+ KTRs, 1,860 KTRs, CTS/ selected and unselected cohort | Increased AMR rates/ activity with increasing risk genotypes (when combined with KLRC2) | No effect on graft survival |
| Diebold M, et al. Am J Transplant. 2023. PMID: 38097018. | 86 DSA+ KTRs/ selected cohort | V allele (FV and VV) associated with MVI | No effect on graft survival or graft function decline |
| Buxeda A, et al. Am J Transplant. 2023. PMID: 36710135. | 221 KTRs (116 controls, 73 AMR and 32 iMVI)/ case control | V allele associated with increased glomerular CD3+ (6.3 ± 6.1 vs 3.4 ± 1.8, P = .051) and CD8+ cell infiltration (3.9 ± 4.9 vs 1.7 ± 1.2, P = 0.05) | NA |
| Bailly E, et al. Am J Transplant. 2024. PMID: 39332679. | 224 KTRs: 118 DSA+ KTR (59 mixed AMR, 59 without rejection), 124 DSA- age and sex matched controls (62 DSA- with TCMR, 62 DSA- without rejection)/ case control | V allele (FV and VV) colocalized with inflammatory and vascular lesions, and with C4d deposition in multiple correspondence analysis | V allele (F/V and V/V) associated with decreased graft survival |
| Diebold M, et al. Transplantation. 2024. PMID: 39402708. | 507 KTRs/ unselected cohort | No association of V/V158 with MVI in whole cohort, borderline significant association with MVI in DSA+ KTR (preformed or *de novo*) | No effect on graft survival or graft function decline |

| | | | |
|---|---|---|---|
| Current study | 1259 KTRs/ unselected cohort; 502 F/F158 (0.40), 577 F/V158 (0.46), 180 V/V158 (0.14) | V/V158 significantly predicted higher rates of chronic active AMR and transplant glomerulopathy<br>The V158 allele significantly and proportionally (F/V < V/V) associated with lower C4d without rejection rates | V/V was a significant and independent predictor of accelerated graft functional decline and graft failure |

**Table S2. STROBE Statement.**

STROBE Statement: Checklist of items that should be included in reports of cohort studies

| | Item No | Recommendation | Page No |
|---|---|---|---|
| Title and abstract | 1 | (*a*) Indicate the study's design with a commonly used term in the title or the abstract | 1, 4 |
| | | (*b*) Provide in the abstract an informative and balanced summary of what was done and what was found | 4 |
| Introduction | | | |
| Background/rationale | 2 | Explain the scientific background and rationale for the investigation being reported | 6-7 |
| Objectives | 3 | State specific objectives, including any prespecified hypotheses | 7 |
| Methods | | | |
| Study design | 4 | Present key elements of study design early in the paper | 8-11 |
| Setting | 5 | Describe the setting, locations, and relevant dates, including periods of recruitment, exposure, follow-up, and data collection | 8-11 |
| Participants | 6 | (*a*) Give the eligibility criteria, and the sources and methods of selection of participants. Describe methods of follow-up | 8-11 |
| | | (*b*) For matched studies, give matching criteria and number of exposed and unexposed | Not applicable |
| Variables | 7 | Clearly define all outcomes, exposures, predictors, potential confounders, and effect modifiers. Give diagnostic criteria, if applicable | 8-11 |
| Data sources/ measurement | 8* | For each variable of interest, give sources of data and details of methods of assessment (measurement). Describe comparability of assessment methods if there is more than one group | 8-11 |
| Bias | 9 | Describe any efforts to address potential sources of bias | 8-11 |
| Study size | 10 | Explain how the study size was arrived at | 8-11; 12 |

| | Item No | Recommendation | Page No |
|---|---|---|---|
| Quantitative variables | 11 | Explain how quantitative variables were handled in the analyses. If applicable, describe which groupings were chosen and why | 8-11 |
| Statistical methods | 12 | (*a*) Describe all statistical methods, including those used to control for confounding | 9-11 |
| | | (*b*) Describe any methods used to examine subgroups and interactions | 9-11 |
| | | (*c*) Explain how missing data were addressed | 9-11 |
| | | (*d*) If applicable, explain how loss to follow-up was addressed | 9-11 |
| | | (*e*) Describe any sensitivity analyses | 9-11 |
| Results | | | |
| Participants | 13* | (a) Report numbers of individuals at each stage of study—eg numbers potentially eligible, examined for eligibility, confirmed eligible, included in the study, completing follow-up, and analysed | 12-15 |
| | | (b) Give reasons for non-participation at each stage | 12-15 |
| | | (c) Consider use of a flow diagram | Not applicable |
| Descriptive data | 14* | (a) Give characteristics of study participants (eg demographic, clinical, social) and information on exposures and potential confounders | 12, Table 1 |
| | | (b) Indicate number of participants with missing data for each variable of interest | 12-15 |
| | | (c) Summarise follow-up time (eg, average and total amount) | 12 |
| Outcome data | 15* | Report numbers of outcome events or summary measures over time | 12-15 |
| Main results | 16 | (*a*) Give unadjusted estimates and, if applicable, confounder-adjusted estimates and their precision (eg, 95% confidence interval). Make clear which confounders were adjusted for and why they were included | 12-15 |
| | | (*b*) Report category boundaries when continuous variables were categorized | 12-15 |

| | Item No | Recommendation | Page No |
|---|---|---|---|
| | | (*c*) If relevant, consider translating estimates of relative risk into absolute risk for a meaningful time period | Not applicable |
| Other analyses | 17 | Report other analyses done—eg analyses of subgroups and interactions, and sensitivity analyses | 9-11; 12-15 |
| Discussion | | | |
| Key results | 18 | Summarise key results with reference to study objectives | 16-18 |
| Limitations | 19 | Discuss limitations of the study, taking into account sources of potential bias or imprecision. Discuss both direction and magnitude of any potential bias | 18 |
| Interpretation | 20 | Give a cautious overall interpretation of results considering objectives, limitations, multiplicity of analyses, results from similar studies, and other relevant evidence | 16-18 |
| Generalisability | 21 | Discuss the generalisability (external validity) of the study results | 16-18 |
| Other information | | | |
| Funding | 22 | Give the source of funding and the role of the funders for the present study and, if applicable, for the original study on which the present article is based | 19 |

*Give information separately for exposed and unexposed groups.

Note: An Explanation and Elaboration article discusses each checklist item and gives methodological background and published examples of transparent reporting. The STROBE checklist is best used in conjunction with this article (freely available on the Web sites of PLoS Medicine at http://www.plosmedicine.org/, Annals of Internal Medicine at http://www.annals.org/, and Epidemiology at http://www.epidem.com/). Information on the STROBE Initiative is available at http://www.strobe-statement.org.

**Table S3. Hardy Weinberg equilibrium.**

| *Variable* | *Value* |
|---|---|
| *Frequency (F)* | 0,628 |
| *Frequency (V)* | 0,372 |
| *Observed VV* | 180 |
| *Observed FV* | 577 |
| *Observed FF* | 502 |
| *Expected VV* | 174,339 |
| *Expected FV* | 588,323 |
| *Expected FF* | 496,339 |
| *Chi-square test* | 0,49 |
| *Chi-square test with continuity correction* | 0,52 |
| *Likelihood-ratio test* | 0,50 |
| *Exact test with selome p-value* | 0,51 |
| *Exact test with dost p-value* | 0,52 |
| *Exact test with mid p-value* | 0,49 |

**Table S4. Results of univariable and multivariable Cox regression analysis assessing the rates of Banff rejection categories according to FCGR3A F158V and transplant-relevant covariables.**

| | | | | *Univariable* | | *Multivariable* | |
|---|---|---|---|---|---|---|---|
| ***Banff rejection category*** | **N()** | **Event(n)** | **Covariable** | **HR with 95%-CI** | **P value** | **HR with 95%-CI** | **P value** |
| ***Borderline TCMR*** | 1229 | 249 | Repeat TX | 0.840 [0.577 - 1.224] | 0,364 | 0.802 [0.535 - 1.201] | 0,284 |
| | 1229 | 249 | Recipient age | 0.974 [0.886 - 1.072] | 0,590 | 0.929 [0.840 - 1.028] | 0,153 |
| | 1229 | 249 | Male recipient | 0.992 [0.768 - 1.282] | 0,954 | 1.018 [0.784 - 1.322] | 0,892 |
| | 1229 | 249 | Donor age | 1.107 [1.012 - 1.212] | **0,026** | 1.092 [0.991 - 1.204] | 0,076 |
| | 1229 | 249 | Male donor | 0.718 [0.560 - 0.921] | **0,009** | 0.761 [0.592 - 0.980] | **0,034** |
| | 1229 | 249 | Pretransplant HLA-DSA | 1.370 [0.906 - 2.071] | 0,136 | 1.401 [0.897 - 2.188] | 0,138 |
| | 1229 | 249 | HLA-mismatch | 1.126 [1.018 - 1.246] | **0,021** | 1.085 [0.978 - 1.203] | 0,125 |
| | 1229 | 249 | F/V158 | 1.075 [0.823 - 1.405] | 0,596 | 1.047 [0.800 - 1.371] | 0,739 |
| | 1229 | 249 | V/V158 | 0.925 [0.620 - 1.379] | 0,702 | 0.937 [0.628 - 1.398] | 0,751 |
| ***TCMR*** | 1227 | 348 | Repeat TX | 0.947 [0.698 - 1.284] | 0,726 | 0.857 [0.616 - 1.192] | 0,359 |
| | 1227 | 348 | Recipient age | 1.012 [0.931 - 1.099] | 0,785 | 0.989 [0.907 - 1.079] | 0,811 |
| | 1227 | 348 | Male recipient | 0.750 [0.607 - 0.928] | **0,008** | 0.778 [0.627 - 0.966] | **0,023** |
| | 1227 | 348 | Donor age | 1.053 [0.977 - 1.134] | 0,180 | 1.027 [0.949 - 1.111] | 0,507 |
| | 1227 | 348 | Male donor | 0.863 [0.700 - 1.065] | 0,171 | 0.903 [0.730 - 1.117] | 0,349 |
| | 1227 | 348 | Pretransplant HLA-DSA | 1.835 [1.329 - 2.534] | **<0.001** | 1.768 [1.247 - 2.508] | **0,001** |
| | 1227 | 348 | HLA-mismatch | 1.161 [1.068 - 1.261] | **<0.001** | 1.140 [1.046 - 1.243] | **0,003** |
| | 1227 | 348 | F/V158 | 0.973 [0.775 - 1.221] | 0,812 | 0.954 [0.759 - 1.198] | 0,684 |
| | 1227 | 348 | V/V158 | 0.941 [0.680 - 1.303] | 0,715 | 0.979 [0.706 - 1.357] | 0,898 |
| ***Chronic active TCMR*** | 1224 | 16 | Repeat TX | 0.401 [0.053 - 3.036] | 0,376 | 0.624 [0.081 - 4.817] | 0,651 |
| | 1224 | 16 | Recipient age | 1.005 [0.691 - 1.462] | 0,980 | 0.902 [0.602 - 1.351] | 0,617 |
| | 1224 | 16 | Male recipient | 1.373 [0.476 - 3.960] | 0,558 | 1.263 [0.436 - 3.663] | 0,667 |
| | 1224 | 16 | Donor age | 1.233 [0.840 - 1.812] | 0,285 | 1.263 [0.828 - 1.928] | 0,279 |
| | 1224 | 16 | Male donor | 1.459 [0.530 - 4.019] | 0,465 | 1.628 [0.584 - 4.542] | 0,352 |
| | 1224 | 16 | Pretransplant HLA-DSA | 0.000 [0.000 - Inf] | 0,997 | 0.000 [0.000 - Inf] | 0,997 |
| | 1224 | 16 | HLA-mismatch | 1.210 [0.811 - 1.806] | 0,350 | 1.182 [0.779 - 1.794] | 0,432 |

| | | | | | | | |
|---|---|---|---|---|---|---|---|
| | 1224 | 16 | F/V158 | 1.208 [0.383 - 3.807] | 0,747 | 1.215 [0.383 - 3.860] | 0,741 |
| | 1224 | 16 | V/V158 | 2.210 [0.593 - 8.234] | 0,237 | 2.276 [0.609 - 8.506] | 0,221 |
| ***DSA- C4d- MVI*** | 1228 | 168 | Repeat TX | 1.813 [1.263 - 2.604] | **0,001** | 1.694 [1.133 - 2.534] | **0,010** |
| | 1228 | 168 | Recipient age | 1.076 [0.954 - 1.213] | 0,236 | 1.040 [0.916 - 1.180] | 0,545 |
| | 1228 | 168 | Male recipient | 0.693 [0.512 - 0.939] | **0,018** | 0.755 [0.553 - 1.030] | 0,076 |
| | 1228 | 168 | Donor age | 1.167 [1.044 - 1.306] | **0,007** | 1.131 [1.004 - 1.275] | **0,043** |
| | 1228 | 168 | Male donor | 0.822 [0.608 - 1.113] | 0,205 | 0.928 [0.683 - 1.261] | 0,634 |
| | 1228 | 168 | Pretransplant HLA-DSA | 2.898 [1.960 - 4.284] | **<0.001** | 2.197 [1.429 - 3.378] | **<0.001** |
| | 1228 | 168 | HLA-mismatch | 1.200 [1.065 - 1.353] | **0,003** | 1.181 [1.043 - 1.338] | **0,009** |
| | 1228 | 168 | F/V158 | 0.904 [0.652 - 1.252] | 0,542 | 0.831 [0.599 - 1.154] | 0,269 |
| | 1228 | 168 | V/V158 | 0.910 [0.568 - 1.457] | 0,694 | 0.939 [0.585 - 1.507] | 0,795 |
| ***C4d without rejection*** | 1225 | 51 | Repeat TX | 1.633 [0.838 - 3.184] | 0,150 | 1.723 [0.821 - 3.619] | 0,151 |
| | 1225 | 51 | Recipient age | 1.063 [0.854 - 1.323] | 0,583 | 1.036 [0.825 - 1.301] | 0,758 |
| | 1225 | 51 | Male recipient | 0.926 [0.527 - 1.626] | 0,789 | 1.013 [0.570 - 1.801] | 0,965 |
| | 1225 | 51 | Donor age | 1.145 [0.935 - 1.402] | 0,191 | 1.100 [0.886 - 1.365] | 0,387 |
| | 1225 | 51 | Male donor | 0.986 [0.569 - 1.708] | 0,959 | 1.099 [0.629 - 1.922] | 0,740 |
| | 1225 | 51 | Pretransplant HLA-DSA | 2.209 [1.038 - 4.699] | **0,040** | 1.924 [0.845 - 4.379] | 0,119 |
| | 1225 | 51 | HLA-mismatch | 1.262 [1.017 - 1.568] | **0,035** | 1.263 [1.004 - 1.589] | **0,047** |
| | 1225 | 51 | F/V158 | 0.514 [0.287 - 0.923] | **0,026** | 0.479 [0.266 - 0.861] | **0,014** |
| | 1225 | 51 | V/V158 | 0.275 [0.084 - 0.902] | **0,033** | 0.276 [0.084 - 0.907] | **0,034** |
| ***Probable AMR*** | 1224 | 28 | Repeat TX | 4.005 [1.875 - 8.552] | **<0.001** | 0.904 [0.358 - 2.281] | 0,830 |
| | 1224 | 28 | Recipient age | 0.872 [0.664 - 1.146] | 0,326 | 0.906 [0.686 - 1.197] | 0,486 |
| | 1224 | 28 | Male recipient | 0.608 [0.290 - 1.279] | 0,190 | 1.146 [0.512 - 2.563] | 0,740 |
| | 1224 | 28 | Donor age | 0.999 [0.769 - 1.297] | 0,994 | 1.080 [0.831 - 1.402] | 0,566 |
| | 1224 | 28 | Male donor | 1.334 [0.625 - 2.848] | 0,457 | 1.529 [0.707 - 3.305] | 0,281 |
| | 1224 | 28 | Pretransplant HLA-DSA | 35.003 [15.314 - 80.005] | **<0.001** | 36.760 [14.228 - 94.978] | **<0.001** |
| | 1224 | 28 | HLA-mismatch | 1.050 [0.779 - 1.417] | 0,748 | 1.006 [0.723 - 1.398] | 0,974 |
| | 1224 | 28 | F/V158 | 2.471 [0.974 - 6.267] | 0,057 | 1.978 [0.772 - 5.067] | 0,155 |
| | 1224 | 28 | V/V158 | 2.341 [0.714 - 7.673] | 0,160 | 2.317 [0.672 - 7.984] | 0,183 |

| | | | | | | | |
|---|---|---|---|---|---|---|---|
| | 1225 | 96 | Repeat TX | 3.637 [2.399 - 5.513] | **<0.001** | 1.327 [0.793 - 2.221] | 0,281 |
| | 1225 | 96 | Recipient age | 0.790 [0.683 - 0.914] | **0,002** | 0.759 [0.652 - 0.882] | **<0.001** |
| | 1225 | 96 | Male recipient | 0.677 [0.453 - 1.012] | 0,057 | 0.945 [0.626 - 1.425] | 0,786 |
| | 1225 | 96 | Donor age | 1.012 [0.880 - 1.163] | 0,871 | 1.136 [0.976 - 1.323] | 0,101 |
| ***Active AMR*** | 1225 | 96 | Male donor | 0.728 [0.487 - 1.088] | 0,121 | 0.748 [0.497 - 1.124] | 0,162 |
| | 1225 | 96 | Pretransplant HLA-DSA | 15.501 [10.352 - 23.213] | **<0.001** | 14.290 [8.892 - 22.965] | **<0.001** |
| | 1225 | 96 | HLA-mismatch | 1.079 [0.920 - 1.264] | 0,350 | 1.042 [0.878 - 1.236] | 0,639 |
| | 1225 | 96 | F/V158 | 0.951 [0.613 - 1.475] | 0,823 | 0.890 [0.573 - 1.384] | 0,606 |
| | 1225 | 96 | V/V158 | 1.200 [0.669 - 2.152] | 0,541 | 1.324 [0.727 - 2.410] | 0,358 |
| | 1224 | 13 | Repeat TX | 1.872 [0.514 - 6.811] | 0,341 | 0.587 [0.110 - 3.137] | 0,534 |
| | 1224 | 13 | Recipient age | 0.623 [0.419 - 0.925] | **0,019** | 0.602 [0.395 - 0.918] | **0,018** |
| | 1224 | 13 | Male recipient | 1.220 [0.397 - 3.747] | 0,729 | 1.496 [0.401 - 5.584] | 0,549 |
| | 1224 | 13 | Donor age | 0.794 [0.548 - 1.151] | 0,224 | 0.973 [0.646 - 1.463] | 0,894 |
| ***Chronic active AMR*** | 1224 | 13 | Male donor | 0.760 [0.255 - 2.264] | 0,623 | 0.893 [0.288 - 2.773] | 0,845 |
| | 1224 | 13 | Pretransplant HLA-DSA | 7.500 [2.255 - 24.937] | **0,001** | 14.056 [2.837 - 69.633] | **0,001** |
| | 1224 | 13 | HLA-mismatch | 1.098 [0.694 - 1.735] | 0,690 | 1.003 [0.596 - 1.690] | 0,990 |
| | 1224 | 13 | F/V158 | 1.791 [0.328 - 9.785] | 0,501 | 1.308 [0.226 - 7.561] | 0,764 |
| | 1224 | 13 | V/V158 | 9.131 [1.894 - 44.028] | **0,006** | 9.912 [1.983 - 49.533] | **0,005** |
| | 1224 | 14 | Repeat TX | 4.819 [1.670 - 13.906] | **0,004** | 0.339 [0.089 - 1.299] | 0,115 |
| | 1224 | 14 | Recipient age | 0.689 [0.474 - 1.002] | 0,051 | 0.648 [0.437 - 0.961] | **0,031** |
| | 1224 | 14 | Male recipient | 0.539 [0.186 - 1.559] | 0,254 | 1.703 [0.473 - 6.124] | 0,415 |
| | 1224 | 14 | Donor age | 1.139 [0.766 - 1.693] | 0,520 | 1.110 [0.721 - 1.707] | 0,636 |
| ***Chronic AMR*** | 1224 | 14 | Male donor | 0.342 [0.107 - 1.090] | 0,070 | 0.550 [0.160 - 1.896] | 0,344 |
| | 1224 | 14 | Pretransplant HLA-DSA | 61.210 [16.824 - 222.704] | **<0.001** | 211.354 [34.783 - 1284.264] | **<0.001** |
| | 1224 | 14 | HLA-mismatch | 0.800 [0.518 - 1.235] | 0,313 | 0.550 [0.300 - 1.009] | 0,054 |
| | 1224 | 14 | F/V158 | 1.040 [0.317 - 3.409] | 0,949 | 0.283 [0.069 - 1.153] | 0,078 |
| | 1224 | 14 | V/V158 | 1.637 [0.391 - 6.854] | 0,500 | 1.185 [0.252 - 5.567] | 0,830 |
| ***No rejection*** | 1248 | 1223 | Repeat TX | 0.865 [0.735 - 1.017] | 0,080 | 0.948 [0.799 - 1.125] | 0,541 |
| | 1248 | 1223 | Recipient age | 1.024 [0.980 - 1.070] | 0,284 | 1.013 [0.967 - 1.062] | 0,591 |

| | | | | | | | |
|---|---|---|---|---|---|---|---|
| | 1248 | 1223 | Male recipient | 1.040 [0.926 - 1.168] | 0,510 | 1.016 [0.903 - 1.144] | 0,790 |
| | 1248 | 1223 | Donor age | 1.033 [0.994 - 1.073] | 0,099 | 1.034 [0.993 - 1.078] | 0,109 |
| | 1248 | 1223 | Male donor | 1.007 [0.900 - 1.127] | 0,901 | 1.003 [0.895 - 1.124] | 0,961 |
| | 1248 | 1223 | Pretransplant HLA-DSA | 0.670 [0.539 - 0.832] | **<0.001** | 0.686 [0.547 - 0.860] | **0,001** |
| | 1248 | 1223 | HLA-mismatch | 0.986 [0.942 - 1.031] | 0,531 | 0.975 [0.931 - 1.021] | 0,283 |
| | 1248 | 1223 | F/V158 | 0.936 [0.830 - 1.057] | 0,289 | 0.946 [0.837 - 1.068] | 0,368 |
| | 1248 | 1223 | V/V158 | 0.900 [0.757 - 1.071] | 0,235 | 0.911 [0.765 - 1.084] | 0,294 |

**Table S5. Results of univariable and multivariable Cox regression analysis assessing the rates of transplant glomerulopathy (cg) according to FCGR3A F158V and transplant-relevant covariables at different grades of cg (presence of lesion [cg=1-3 versus 0], medium grade lesion [cg=2,3 versus 0, 1 or high-grade lesion [cg=3 versus 0, 1, 2]).**

| | | *Cg = 1, 2, 3 vs 0* | | | *Cg = 2, 3 vs 0, 1* | | | *Cg = 3 vs 0, 1, 2* | | |
|---|---|---|---|---|---|---|---|---|---|---|
| *Univariable model* | | | | | | | | | | |
| ***N()*** | **Covariable** | **Event(n)** | **HR [95%-CI]** | **P value** | **Events** | **HR [95%-CI]** | **P value** | **Events** | **HR [95%-CI]** | **P value** |
| | N() | **1246** | | | **1245** | | | **1245** | | |
| *1246* | Repeat TX | 119 | 1.787 [1.157 - 2.761] | **0,009** | 45 | 2.372 [1.225 - 4.595] | **0,010** | 27 | 2.700 [1.181 - 6.173] | **0,019** |
| *1246* | Recipient age | 119 | 1.055 [0.916 - 1.215] | 0,456 | 45 | 0.908 [0.730 - 1.128] | 0,382 | 27 | 0.948 [0.714 - 1.259] | 0,713 |
| *1246* | Male recipient | 119 | 0.809 [0.563 - 1.162] | 0,252 | 45 | 0.761 [0.423 - 1.369] | 0,362 | 27 | 0.645 [0.303 - 1.376] | 0,256 |
| *1246* | Donor age | 119 | 1.087 [0.952 - 1.240] | 0,217 | 45 | 1.032 [0.836 - 1.273] | 0,772 | 27 | 1.148 [0.864 - 1.526] | 0,341 |
| *1246* | Male donor | 119 | 1.024 [0.714 - 1.470] | 0,896 | 45 | 1.299 [0.715 - 2.360] | 0,390 | 27 | 1.084 [0.507 - 2.316] | 0,836 |
| *1246* | Pretransplant HLA-DSA | 119 | 2.780 [1.714 - 4.509] | **<0.001** | 45 | 6.608 [3.486 - 12.527] | **<0.001** | 27 | 10.563 [4.818 - 23.159] | **<0.001** |
| *1246* | HLA-mismatch | 119 | 1.013 [0.873 - 1.176] | 0,860 | 45 | 1.086 [0.852 - 1.383] | 0,506 | 27 | 0.843 [0.617 - 1.153] | 0,286 |
| *1246* | F/V158 | 119 | 1.115 [0.743 - 1.671] | 0,600 | 45 | 1.270 [0.641 - 2.514] | 0,494 | 27 | 1.526 [0.600 - 3.877] | 0,375 |
| *1246* | V/V158 | 119 | 1.666 [1.009 - 2.751] | **0,046** | 45 | 2.262 [1.027 - 4.984] | **0,043** | 27 | 3.205 [1.162 - 8.840] | **0,024** |
| *Multivariable model* | | | | | | | | | | |
| ***N()*** | **Covariable** | **Event(n)** | **HR [95%-CI]** | **P value** | **Events** | **HR [95%-CI]** | **P value** | **Events** | **HR [95%-CI]** | **P value** |
| *1246* | Repeat TX | 119 | 1.458 [0.887 - 2.395] | 0,137 | 45 | 1.154 [0.515 - 2.588] | 0,727 | 27 | 0.825 [0.293 - 2.325] | 0,717 |
| *1246* | Recipient age | 119 | 1.061 [0.915 - 1.229] | 0,434 | 45 | 0.914 [0.730 - 1.144] | 0,432 | 27 | 0.918 [0.687 - 1.226] | 0,561 |
| *1246* | Male recipient | 119 | 0.892 [0.616 - 1.293] | 0,548 | 45 | 0.968 [0.524 - 1.786] | 0,916 | 27 | 0.947 [0.423 - 2.122] | 0,895 |
| *1246* | Donor age | 119 | 1.090 [0.950 - 1.252] | 0,219 | 45 | 1.071 [0.863 - 1.329] | 0,533 | 27 | 1.198 [0.896 - 1.602] | 0,222 |
| *1246* | Male donor | 119 | 1.100 [0.763 - 1.586] | 0,609 | 45 | 1.482 [0.810 - 2.712] | 0,202 | 27 | 1.314 [0.604 - 2.856] | 0,491 |
| *1246* | Pretransplant HLA-DSA | 119 | 2.361 [1.360 - 4.097] | **0,002** | 45 | 6.442 [2.931 - 14.159] | **<0.001** | 27 | 12.690 [4.593 - 35.065] | **<0.001** |
| *1246* | HLA-mismatch | 119 | 1.004 [0.861 - 1.170] | 0,959 | 45 | 1.090 [0.841 - 1.412] | 0,514 | 27 | 0.789 [0.561 - 1.110] | 0,174 |
| *1246* | F/V158 | 119 | 1.042 [0.693 - 1.569] | 0,842 | 45 | 1.027 [0.511 - 2.064] | 0,940 | 27 | 1.092 [0.417 - 2.859] | 0,858 |
| *1246* | V/V158 | 119 | 1.676 [1.014 - 2.771] | **0,044** | 45 | 2.233 [1.008 - 4.947] | **0,048** | 27 | 3.111 [1.115 - 8.685] | **0,030** |

**Table S6. Cumulative incidence of C4d without rejection, probable AMR and full AMR (active AMR, chronic active AMR and chronic AMR) at first biopsy considering these phenotypes as competing risks.**

| | *Number at risk (n)* | *Events (n)* | *Cumulative incidence* | *Overall P value (Gray's test)* |
|---|---|---|---|---|
| ***C4d without rejection*** | | | | |
| *F/F158* | 495 | 23 | 0.057 | |
| *F/V158* | 571 | 14 | 0.027 | |
| *V/V158* | 178 | 2 | 0.014 | **0.034** |
| *Total* | 1244 | 39 | | |
| ***Probable AMR*** | | | | |
| *F/F158* | 495 | 5 | 0.012 | |
| *F/V158* | 571 | 12 | 0.047 | |
| *V/V158* | 178 | 4 | 0.052 | 0.3 |
| *Total* | 1244 | 21 | | |
| ***Full AMR*** | | | | |
| *F/F158* | 495 | 35 | 0.133 | |
| *F/V158* | 571 | 38 | 0.130 | |
| *V/V158* | 178 | 18 | 0.214 | 0.29 |
| *Total* | 1244 | 91 | | |

**Table S7. Results of univariable linear mixed models assessing the association of graft functional decline with FCGR3A F158V genotype and transplant-relevant covariables in the whole cohort, rejection cohort and AMR/MVI cohort.**

| *Term* | *Estimate [95%-CI]* | *Adj.est.* | *P value* | *Estimate [95%-CI]* | *Adj.est.* | *P value* | *Estimate [95%-CI]* | *Adj.est.* | *P value* |
|---|---|---|---|---|---|---|---|---|---|
| | ***Whole cohort 69.354 eGFR measurements*** | | | ***Rejection cohort 28.034 eGFR measurements*** | | | ***AMR/ MVI cohort 15.570 measurements*** | | |
| *Intercept* | 51.622 [49.812, 53.431] | | <0.001 | 47.788 [44.435, 51.141] | | <0.001 | 44.043 [39.454, 48.631] | | <0.001 |
| *F/V158* | 1.141 [-1.329, 3.611] | | 0,365 | 1.51 [-2.973, 5.992] | | 0,508 | 5.615 [-0.609, 11.84] | | 0,077 |
| *V/V158* | 1.865 [-1.661, 5.39] | | 0,300 | 4.655 [-2.014, 11.323] | | 0,171 | 2.512 [-6.46, 11.483] | | 0,582 |
| *Time* | -0.736 [-1.186, -0.287] | -0,736 | 0,001 | -1.043 [-1.803, -0.282] | -1,043 | 0,007 | -1.215 [-2.146, -0.283] | -1,215 | 0,011 |
| *F/V158: time* | -0.556 [-1.169, 0.057] | -1,293 | 0,075 | -0.905 [-1.947, 0.138] | -1,947 | 0,089 | -2.034 [-3.319, -0.749] | -3,248 | **0,002** |
| *V/V158: time* | -0.944 [-1.817, -0.071] | -1,680 | **0,034** | -2.15 [-3.694, -0.606] | -3,193 | **0,007** | -2.549 [-4.391, -0.707] | -3,763 | **0,007** |
| *Intercept* | 51.977 [50.745, 53.208] | | <0.001 | 48.963 [46.472, 51.455] | | <0.001 | 46.385 [42.753, 50.016] | | <0.001 |
| *Repeat TX* | 3.03 [-0.203, 6.264] | | 0,066 | 1.034 [-4.39, 6.459] | | 0,708 | 2.453 [-4.163, 9.07] | | 0,466 |
| *Time* | -0.927 [-1.231, -0.623] | -0,927 | <0.001 | -1.384 [-1.92, -0.848] | -1,384 | <0.001 | -2.014 [-2.711, -1.316] | -2,014 | <0.001 |
| *Repeat TX: time* | -1.409 [-2.215, -0.603] | -2,336 | **0,001** | -1.913 [-3.162, -0.664] | -3,297 | **0,003** | -1.871 [-3.248, -0.494] | -3,884 | **0,008** |
| *Intercept* | 66.175 [61.331, 71.019] | | <0.001 | 61.361 [52.927, 69.795] | | <0.001 | 58.381 [47.112, 69.651] | | <0.001 |
| *Recipient age* | -2.563 [-3.441, -1.686] | | <0.001 | -2.296 [-3.808, -0.783] | | 0,003 | -2.19 [-4.239, -0.141] | | 0,036 |
| *Time* | -2.663 [-3.873, -1.453] | -2,663 | <0.001 | -5.167 [-7.124, -3.211] | -5,167 | <0.001 | -6.432 [-8.766, -4.098] | -6,432 | <0.001 |
| *Recipient age: time* | 0.288 [0.067, 0.508] | -2,375 | **0,011** | 0.641 [0.286, 0.997] | -4,526 | **<0.001** | 0.751 [0.32, 1.182] | -5,681 | **0,001** |
| *Intercept* | 52.523 [50.645, 54.401] | | <0.001 | 48.499 [45.214, 51.784] | | <0.001 | 46.208 [41.656, 50.761] | | <0.001 |
| *Male recpient* | -0.179 [-2.543, 2.185] | | 0,882 | 1.052 [-3.142, 5.245] | | 0,622 | 1.277 [-4.513, 7.066] | | 0,664 |
| *Time* | -1.684 [-2.148, -1.221] | -1,684 | <0.001 | -1.922 [-2.657, -1.188] | -1,922 | <0.001 | -2.71 [-3.63, -1.79] | -2,710 | <0.001 |
| *Male recipient: time* | 0.885 [0.301, 1.469] | -0,800 | **0,003** | 0.322 [-0.662, 1.306] | -1,600 | 0,520 | 0.367 [-0.861, 1.594] | -2,343 | 0,556 |
| *Intercept* | 65.911 [59.361, 72.461] | | <0.001 | 59.07 [47.49, 70.651] | | <0.001 | 63.664 [47.328, 80.001] | | <0.001 |
| *Recipient BMI* | -0.53 [-0.784, -0.277] | | <0.001 | -0.385 [-0.823, 0.053] | | 0,084 | -0.641 [-1.253, -0.029] | | 0,040 |
| *Time* | -2.791 [-4.434, -1.149] | -2,791 | 0,001 | -3.955 [-6.7, -1.21] | -3,955 | 0,005 | -4.894 [-8.41, -1.378] | -4,894 | 0,007 |
| *Recipient BMI:time* | 0.066 [0.002, 0.129] | -2,726 | **0,044** | 0.086 [-0.019, 0.19] | -3,869 | 0,108 | 0.092 [-0.041, 0.226] | -4,802 | 0,175 |
| *Intercept* | 52.307 [51.119, 53.495] | | <0.001 | 48.628 [46.162, 51.095] | | <0.001 | 45.05 [41.329, 48.771] | | <0.001 |
| *Pre-TX HLA-DSA* | 1.426 [-2.768, 5.619] | | 0,505 | 3.509 [-2.247, 9.264] | | 0,231 | 6.731 [0.287, 13.175] | | 0,041 |

| | | | | | | | | | |
|---|---|---|---|---|---|---|---|---|---|
| *Time* | -0.954 [-1.246, -0.662] | -0,954 | <0.001 | -1.368 [-1.888, -0.848] | -1,368 | <0.001 | -1.962 [-2.657, -1.267] | -1,962 | <0.001 |
| *Pre-TX HLA-DSA:time* | -2.203 [-3.244, -1.161] | -3,157 | **<0.001** | -2.455 [-3.795, -1.115] | -3,822 | **<0.001** | -2.018 [-3.364, -0.672] | -3,980 | **0,004** |
| *Intercept* | 50.795 [48.006, 53.583] | | <0.001 | 47.933 [42.426, 53.44] | | <0.001 | 44.601 [37.488, 51.713] | | <0.001 |
| *CIT* | 0.616 [-0.354, 1.586] | | 0,213 | 0.425 [-1.438, 2.288] | | 0,654 | 0.845 [-1.517, 3.208] | | 0,481 |
| *Time* | -0.711 [-1.402, -0.02] | -0,711 | 0,044 | -0.972 [-2.244, 0.299] | -0,972 | 0,133 | -1.663 [-3.117, -0.209] | -1,663 | 0,025 |
| *CIT:time* | -0.158 [-0.398, 0.081] | -0,869 | 0,195 | -0.285 [-0.718, 0.148] | -1,257 | 0,197 | -0.311 [-0.797, 0.175] | -1,974 | 0,209 |
| *Intercept* | 81.77 [78.154, 85.386] | | <0.001 | 81.364 [74.386, 88.342] | | <0.001 | 84.612 [74.781, 94.443] | | <0.001 |
| *Donor age* | -6.088 [-6.808, -5.368] | | <0.001 | -6.485 [-7.828, -5.142] | | <0.001 | -7.528 [-9.394, -5.662] | | <0.001 |
| *Time* | -2.026 [-3.022, -1.029] | -2,026 | <0.001 | -3.881 [-5.636, -2.125] | -3,881 | <0.001 | -7.055 [-9.267, -4.844] | -7,055 | <0.001 |
| *Donor age: time* | 0.188 [-0.011, 0.388] | -1,837 | 0,064 | 0.434 [0.09, 0.779] | -3,446 | **0,014** | 0.917 [0.485, 1.349] | -6,138 | **<0.001** |
| *Intercept* | 49.677 [48.016, 51.337] | | <0.001 | 46.109 [43.066, 49.151] | | <0.001 | 44.974 [40.809, 49.139] | | <0.001 |
| *Male donor* | 5.111 [2.842, 7.381] | | <0.001 | 5.996 [1.87, 10.122] | | 0,004 | 4.058 [-1.683, 9.798] | | 0,165 |
| *Time* | -1.051 [-1.466, -0.635] | -1,051 | <0.001 | -1.319 [-2.015, -0.623] | -1,319 | <0.001 | -2.164 [-3.011, -1.317] | -2,164 | <0.001 |
| *Male donor: time* | -0.144 [-0.712, 0.424] | -1,195 | 0,619 | -0.839 [-1.813, 0.136] | -2,158 | 0,091 | -0.709 [-1.928, 0.51] | -2,873 | 0,252 |
| *Intercept* | 52.128 [50.805, 53.451] | | <0.001 | 48.9 [46.341, 51.459] | | <0.001 | 46.333 [42.711, 49.955] | | <0.001 |
| *DCD* | 1.191 [-1.768, 4.149] | | 0,430 | -0.548 [-6.134, 5.038] | | 0,847 | 1.623 [-6.355, 9.601] | | 0,689 |
| *LD* | 0.898 [-3.597, 5.393] | | 0,695 | 4.607 [-4.243, 13.457] | | 0,307 | 3.198 [-7.214, 13.61] | | 0,545 |
| *Time* | -1.164 [-1.493, -0.835] | -1,164 | <0.001 | -1.785 [-2.344, -1.227] | -1,785 | <0.001 | -2.596 [-3.303, -1.889] | -2,596 | <0.001 |
| *DCD: time* | 0.153 [-0.584, 0.89] | -1,011 | 0,684 | 0.405 [-0.93, 1.74] | -1,381 | 0,551 | 0.105 [-1.62, 1.831] | -2,490 | 0,904 |
| *LD: time* | 0.105 [-0.995, 1.206] | -1,059 | 0,851 | -0.398 [-2.441, 1.645] | -2,184 | 0,702 | 0.759 [-1.377, 2.895] | -1,837 | 0,484 |
| *Intercept* | 56.535 [53.956, 59.114] | | <0.001 | 52.429 [47.365, 57.493] | | <0.001 | 50.069 [43.181, 56.957] | | <0.001 |
| *HLA mismatch* | -1.64 [-2.561, -0.719] | | <0.001 | -1.258 [-2.947, 0.431] | | 0,144 | -1.17 [-3.478, 1.137] | | 0,319 |
| *Time* | -1.745 [-2.382, -1.108] | -1,745 | <0.001 | -2.688 [-3.846, -1.531] | -2,688 | <0.001 | -4.231 [-5.669, -2.793] | -4,231 | <0.001 |
| *HLA mismatch: time* | 0.247 [0.019, 0.475] | -1,498 | **0,034** | 0.356 [-0.039, 0.751] | -2,333 | 0,077 | 0.641 [0.157, 1.124] | -3,590 | **0,010** |

Linear regression for the rejection and AMR/MVI cohort was corrected for time to first rejection. Significant values are represented in **bold**. Adjusted estimates were calculated as the sum of the main effect of time and the slope estimate for each covariable.

**Table S8. Results of multivariable linear mixed models assessing the association of graft functional decline with FCGR3A F158V genotype and transplant-relevant covariables in the whole cohort, rejection cohort and AMR/MVI cohort.**

| *Covariable* | *Estimate [95%-CI]* | *P value* | *Estimate [95%-CI]* | *Adj.est.* | *P value* |
|---|---|---|---|---|---|
| | ***Whole cohort (69.354 eGFR values)*** | | | | |
| | ***Main effect*** | | ***Slope with time*** | | |
| *Intercept* | 87.6 [78.924, 96.276] | 0,000 | | | |
| *Time* | -3.439 [-5.812, -1.065] | 0,005 | | | |
| *F/V158* | 1.876 [-0.347, 4.099] | 0,098 | -0.568 [-1.176, 0.039] | -4,007 | 0,066 |
| *V/V158* | 0.869 [-2.298, 4.037] | 0,590 | -0.946 [-1.809, -0.082] | -4,384 | **0,032** |
| *Repeat TX* | -0.586 [-3.719, 2.547] | 0,714 | -0.662 [-1.523, 0.2] | -4,100 | 0,132 |
| *Recipient age* | -0.322 [-1.201, 0.556] | 0,472 | 0.207 [-0.032, 0.446] | -3,232 | 0,090 |
| *Male recipient* | 0.262 [-1.876, 2.401] | 0,810 | 0.748 [0.165, 1.331] | -2,690 | **0,012** |
| *Recipient BMI* | -0.357 [-0.592, -0.121] | 0,003 | 0.046 [-0.019, 0.11] | -3,393 | 0,168 |
| *Pretransplant HLA-DSA* | 1.032 [-2.976, 5.04] | 0,614 | -1.7 [-2.801, -0.599] | -5,139 | **0,003** |
| *CIT* | 0.474 [-0.648, 1.596] | 0,407 | -0.183 [-0.49, 0.124] | -3,622 | 0,242 |
| *Donor age* | -5.769 [-6.543, -4.994] | 0,000 | 0.06 [-0.152, 0.272] | -3,379 | 0,579 |
| *Male donor* | 2.747 [0.65, 4.844] | 0,010 | -0.041 [-0.615, 0.532] | -3,480 | 0,888 |
| *DCD* | 0.092 [-2.603, 2.788] | 0,946 | 0.084 [-0.65, 0.819] | -3,355 | 0,822 |
| *LD* | 3.14 [-2.212, 8.491] | 0,250 | -0.28 [-1.726, 1.167] | -3,718 | 0,704 |
| *HLA mismatch* | -0.272 [-1.152, 0.608] | 0,544 | 0.173 [-0.066, 0.412] | -3,266 | 0,156 |
| | ***Rejection cohort (28.034 eGFR values)*** | | | | |
| | ***Main effect*** | | ***Slope with time*** | | |
| *Intercept* | 78.756 [62.613, 94.899] | <0.001 | | | |
| *Time* | -4.271 [-8.348, -0.193] | 0,040 | | | |
| *F/V158* | 0.446 [-3.696, 4.589] | 0,832 | -0.803 [-1.842, 0.235] | -5,074 | 0,129 |
| *V/V158* | 2.784 [-3.359, 8.928] | 0,373 | -1.89 [-3.419, -0.36] | -6,160 | **0,016** |
| *Repeat TX* | -1.342 [-7.088, 4.404] | 0,646 | -0.659 [-2.091, 0.772] | -4,930 | 0,365 |
| *Recipient age* | 0.574 [-1.018, 2.166] | 0,479 | 0.417 [0.019, 0.816] | -3,853 | **0,040** |
| *Male recipient* | -0.05 [-3.959, 3.86] | 0,980 | 0.274 [-0.705, 1.253] | -3,997 | 0,582 |

| | | | | | |
|---|---|---|---|---|---|
| *Recipient BMI* | -0.314 [-0.722, 0.094] | 0,131 | 0.073 [-0.031, 0.177] | -4,198 | 0,169 |
| *Pretransplant HLA-DSA* | 3.026 [-2.98, 9.032] | 0,323 | -1.681 [-3.199, -0.162] | -5,952 | **0,030** |
| *CIT* | 1.414 [-0.774, 3.601] | 0,205 | -0.518 [-1.065, 0.029] | -4,789 | 0,063 |
| *Donor age* | -6.532 [-8.062, -5.001] | <0.001 | 0.153 [-0.232, 0.537] | -4,118 | 0,435 |
| *Male donor* | 3.176 [-0.774, 7.127] | 0,115 | -0.68 [-1.668, 0.308] | -4,951 | 0,177 |
| *DCD* | -2.284 [-7.494, 2.926] | 0,389 | 0.388 [-0.929, 1.705] | -3,883 | 0,562 |
| *LD* | 7.124 [-3.491, 17.739] | 0,188 | -1.212 [-3.835, 1.411] | -5,483 | 0,364 |
| *HLA mismatch* | 0.639 [-1.077, 2.355] | 0,465 | 0.104 [-0.326, 0.533] | -4,167 | 0,635 |
| | ***AMR/MVI cohort (15.570 eGFR values)*** | | | | |
| | ***Main effect*** | | ***Slope with time*** | | |
| *Intercept* | 86.082 [63.779, 108.386] | <0.001 | | | |
| *Time* | -7.961 [-13.011, -2.912] | 0,002 | | | |
| *F/V158* | 3.14 [-2.607, 8.886] | 0,283 | -1.645 [-2.937, -0.354] | -9,607 | **0,013** |
| *V/V158* | -0.394 [-8.502, 7.715] | 0,924 | -2.076 [-3.9, -0.252] | -10,038 | **0,026** |
| *Repeat TX* | -2.203 [-9.45, 5.044] | 0,550 | -0.583 [-2.232, 1.067] | -8,544 | 0,486 |
| *Recipient age* | 0.736 [-1.433, 2.905] | 0,504 | 0.434 [-0.053, 0.922] | -7,527 | 0,080 |
| *Male recipient* | -0.675 [-5.987, 4.636] | 0,802 | 0.596 [-0.598, 1.79] | -7,365 | 0,325 |
| *Recipient BMI* | -0.501 [-1.06, 0.059] | 0,079 | 0.078 [-0.052, 0.209] | -7,883 | 0,238 |
| *Pretransplant HLA-DSA* | 5.479 [-1.524, 12.482] | 0,124 | -1.027 [-2.607, 0.554] | -8,988 | 0,201 |
| *CIT* | 1.676 [-1.253, 4.606] | 0,260 | -0.383 [-1.029, 0.264] | -8,344 | 0,244 |
| *Donor age* | -7.791 [-9.94, -5.641] | <0.001 | 0.581 [0.103, 1.059] | -7,381 | **0,018** |
| *Male donor* | 0.409 [-4.917, 5.735] | 0,880 | -0.345 [-1.537, 0.848] | -8,306 | 0,569 |
| *DCD* | 4.086 [-3.201, 11.372] | 0,270 | -0.01 [-1.672, 1.653] | -7,971 | 0,991 |
| *LD* | 6.189 [-7.491, 19.869] | 0,373 | -0.209 [-3.205, 2.787] | -8,171 | 0,890 |
| *HLA mismatch* | 0.439 [-1.962, 2.84] | 0,719 | 0.233 [-0.311, 0.777] | -7,729 | 0,399 |

Multivariable linear mixed regression models were corrected for transplant-relevant covariables (repeat transplantation [yes/ no], recipient age [/10 years], recipient sex [male or female], recipient BMI [/kg/m²], pretransplant HLA DSA status [present/ absent], cold ischemia time [/hours], donor age [/10 years], donor sex [male or female], donor type [living donation, donation after brain death, donation after cardiac death], HLA mismatch [/mismatch at HLA-A/B/DR]. For the rejection and AMR/MVI cohort, regression models were also corrected for time to first rejection.

**Table S9. Results of multivariable Cox and competing risk regression models assessing the association between FCGR3A F158V genotype and transplant-relevant covariables with graft failure in the whole cohort, rejection cohort and AMR/MVI cohort.**

| | | ***Whole cohort*** *TX=1259, events=205* | | ***Rejection cohort*** *TX=459, events=113* | | ***AMR/ MVI cohort*** *TX=239, events=70* | |
|---|---|---|---|---|---|---|---|
| | **Covariable** | **HR [95%-CI]** | **P value** | **HR [95%-CI]** | **P value** | **HR [95%-CI]** | **P value** |
| *Cox regression* | *Repeat TX* | 1.271 [0.862 - 1.874] | 0,226 | 1.416 [0.853 - 2.350] | 0,179 | 1.385 [0.739 - 2.596] | 0,309 |
| | *Recipient age (/10 years)* | 0.884 [0.786 - 0.993] | **0,038** | 0.822 [0.705 - 0.958] | **0,012** | 0.829 [0.686 - 1.002] | **0,053** |
| | *Male recipient* | 0.798 [0.603 - 1.056] | 0,115 | 0.845 [0.578 - 1.236] | 0,386 | 0.582 [0.353 - 0.960] | **0,034** |
| | *Recipient BMI (/[kg/m²])* | 0.991 [0.960 - 1.022] | 0,570 | 1.010 [0.970 - 1.052] | 0,639 | 0.997 [0.945 - 1.052] | 0,911 |
| | *Cold ischemia time (/5 hours)* | 1.159 [1.028 - 1.307] | **0,016** | 1.090 [0.918 - 1.294] | 0,325 | 1.070 [0.865 - 1.325] | 0,532 |
| | *Donor age (/10 years)* | 1.207 [1.080 - 1.350] | **0,001** | 1.094 [0.935 - 1.280] | 0,261 | 0.925 [0.766 - 1.116] | 0,415 |
| | *Male donor* | 1.121 [0.849 - 1.482] | 0,420 | 1.026 [0.698 - 1.507] | 0,897 | 1.318 [0.800 - 2.173] | 0,278 |
| | *Pretransplant HLA-DSA* | 2.198 [1.454 - 3.323] | **0,000** | 1.587 [0.947 - 2.658] | 0,079 | 1.445 [0.766 - 2.724] | 0,256 |
| | *HLA mismatch (A/B/DR)* | 1.110 [0.985 - 1.251] | 0,087 | 1.168 [0.980 - 1.393] | 0,083 | 0.965 [0.767 - 1.213] | 0,758 |
| | *F/V158* | 1.108 [0.813 - 1.511] | 0,516 | 1.294 [0.844 - 1.984] | 0,237 | 1.371 [0.769 - 2.444] | 0,284 |
| | *V/V158* | 1.546 [1.045 - 2.287] | **0,029** | 1.877 [1.098 - 3.209] | **0,021** | 1.966 [0.988 - 3.911] | **0,054** |
| *Competing risk regression* | *Repeat TX* | 1.203 [0.807 - 1.794] | 0,360 | 1.452 [0.858 - 2.458] | 0,160 | 1.409 [0.728 - 2.725] | 0,310 |
| | *Recipient age (/10 years)* | 0.818 [0.728 - 0.918] | **0,001** | 0.764 [0.654 - 0.892] | **0,001** | 0.786 [0.649 - 0.953] | **0,014** |
| | *Male recipient* | 0.795 [0.604 - 1.047] | 0,100 | 0.831 [0.570 - 1.211] | 0,340 | 0.593 [0.360 - 0.979] | **0,041** |
| | *Recipient BMI (/[kg/m²])* | 0.991 [0.959 - 1.024] | 0,590 | 1.012 [0.972 - 1.053] | 0,570 | 0.995 [0.941 - 1.052] | 0,860 |
| | *Cold ischemia time (/5 hours)* | 1.171 [1.046 - 1.311] | **0,006** | 1.100 [0.943 - 1.283] | 0,220 | 1.072 [0.894 - 1.286] | 0,450 |
| | *Donor age (/10 years)* | 1.195 [1.061 - 1.346] | **0,003** | 1.075 [0.911 - 1.269] | 0,390 | 0.924 [0.766 - 1.116] | 0,410 |
| | *Male donor* | 1.109 [0.838 - 1.469] | 0,470 | 1.063 [0.726 - 1.557] | 0,750 | 1.401 [0.864 - 2.274] | 0,170 |
| | *Pretransplant HLA-DSA* | 2.172 [1.432 - 3.293] | **0,000** | 1.490 [0.887 - 2.503] | 0,130 | 1.401 [0.725 - 2.705] | 0,320 |
| | *HLA mismatch (A/B/DR)* | 1.091 [0.951 - 1.252] | 0,210 | 1.180 [0.977 - 1.425] | 0,086 | 0.966 [0.755 - 1.237] | 0,790 |
| | *F/V158* | 1.127 [0.826 - 1.538] | 0,450 | 1.337 [0.873 - 2.046] | 0,180 | 1.428 [0.786 - 2.597] | 0,240 |
| | *V/V158* | 1.583 [1.078 - 2.325] | **0,019** | 1.815 [1.068 - 3.086] | **0,028** | 2.024 [1.060 - 3.866] | **0,033** |

Multivariable models were corrected for transplant-relevant covariables (repeat transplantation [yes/ no], recipient age [/10 years], recipient sex [male or female], recipient BMI [/kg/m²], pretransplant HLA DSA status [present/ absent], cold ischemia time [/5 hours], donor age [/10 years], donor sex [male or female], donor type [living donation, donation after brain death, donation after cardiac death], HLA mismatch [/mismatch at HLA-A/B/DR]. Regression models for the rejection and AMR/ MVI cohort were corrected for time to first rejection. Significant values are represented in **bold**.

**Figure S1. Cumulative incidence of Banff rejection categories stratified by *FCGR3A* F158V genotype**

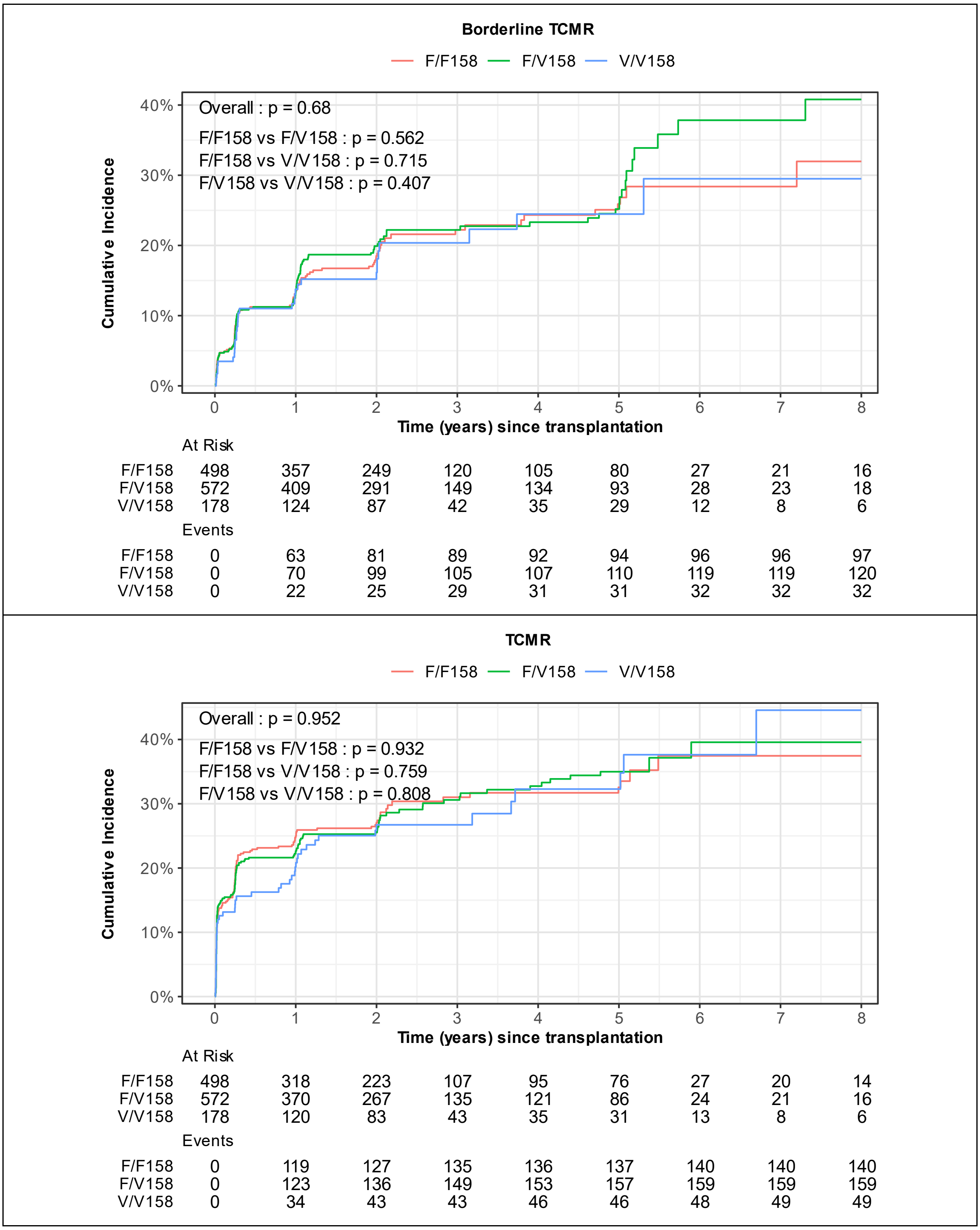

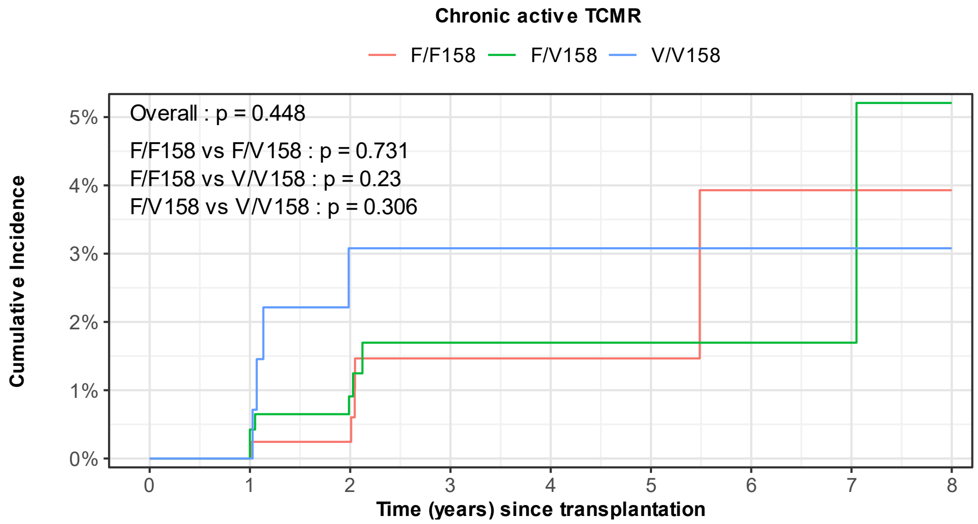


At Risk

| | 0 | 1 | 2 | 3 | 4 | 5 | 6 | 7 | 8 |
|---|---|---|---|---|---|---|---|---|---|
| F/F158 | 498 | 410 | 295 | 150 | 133 | 104 | 37 | 28 | 21 |
| F/V158 | 572 | 468 | 351 | 179 | 161 | 115 | 36 | 28 | 20 |
| V/V158 | 178 | 143 | 102 | 54 | 46 | 39 | 15 | 10 | 8 |

Events

| | 0 | 1 | 2 | 3 | 4 | 5 | 6 | 7 | 8 |
|---|---|---|---|---|---|---|---|---|---|
| F/F158 | 0 | 0 | 1 | 4 | 4 | 4 | 5 | 5 | 5 |
| F/V158 | 0 | 2 | 4 | 6 | 6 | 6 | 6 | 6 | 7 |
| V/V158 | 0 | 0 | 4 | 4 | 4 | 4 | 4 | 4 | 4 |

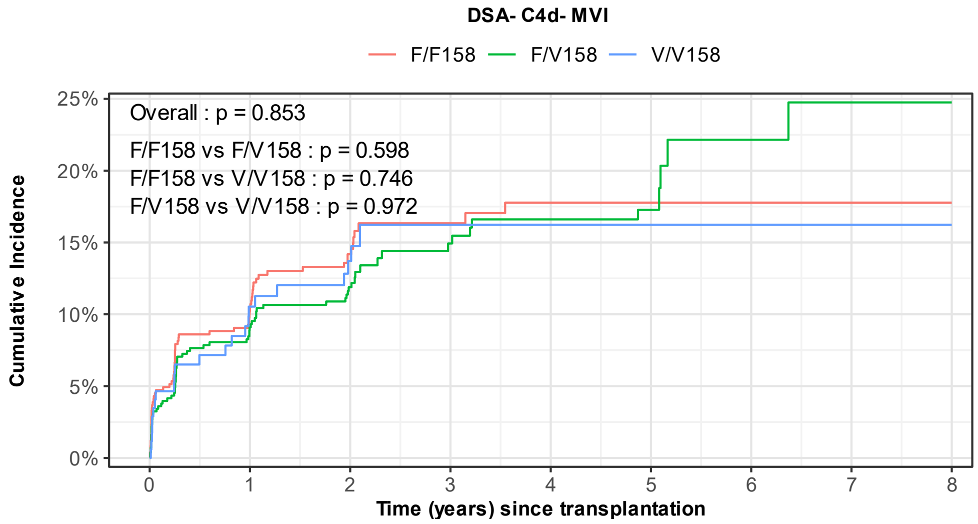


At Risk

| | 0 | 1 | 2 | 3 | 4 | 5 | 6 | 7 | 8 |
|---|---|---|---|---|---|---|---|---|---|
| F/F158 | 498 | 373 | 252 | 124 | 108 | 83 | 29 | 20 | 14 |
| F/V158 | 572 | 431 | 315 | 158 | 138 | 96 | 31 | 26 | 18 |
| V/V158 | 178 | 129 | 86 | 47 | 41 | 34 | 14 | 9 | 7 |

Events

| | 0 | 1 | 2 | 3 | 4 | 5 | 6 | 7 | 8 |
|---|---|---|---|---|---|---|---|---|---|
| F/F158 | 0 | 49 | 63 | 68 | 70 | 70 | 70 | 70 | 70 |
| F/V158 | 0 | 48 | 60 | 67 | 70 | 71 | 74 | 75 | 75 |
| V/V158 | 0 | 17 | 21 | 23 | 23 | 23 | 23 | 23 | 23 |

**C4d without rejection**

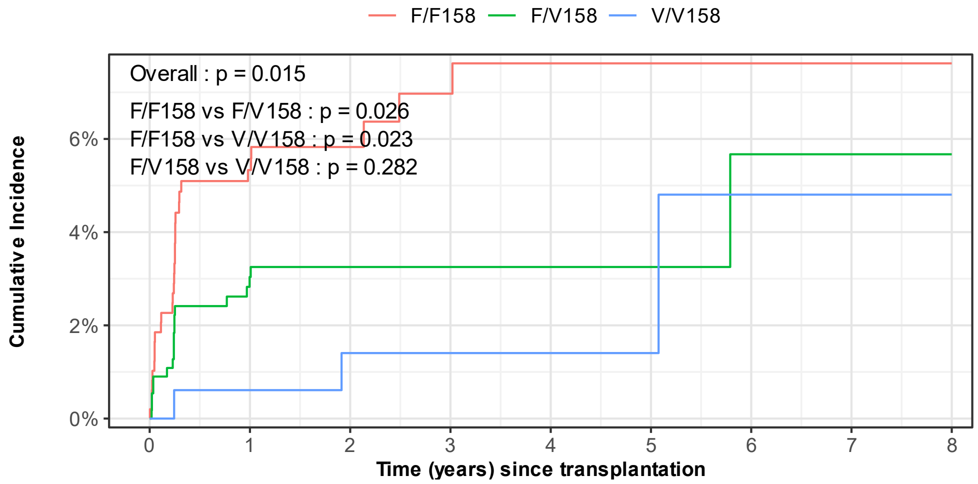


At Risk

| | 0 | 1 | 2 | 3 | 4 | 5 | 6 | 7 | 8 |
|---|---|---|---|---|---|---|---|---|---|
| F/F158 | 498 | 389 | 281 | 146 | 128 | 99 | 36 | 27 | 20 |
| F/V158 | 572 | 455 | 345 | 177 | 159 | 113 | 36 | 28 | 20 |
| V/V158 | 178 | 142 | 101 | 53 | 45 | 39 | 15 | 10 | 8 |

Events

| | 0 | 1 | 2 | 3 | 4 | 5 | 6 | 7 | 8 |
|---|---|---|---|---|---|---|---|---|---|
| F/F158 | 0 | 25 | 27 | 29 | 30 | 30 | 30 | 30 | 30 |
| F/V158 | 0 | 16 | 17 | 17 | 17 | 17 | 18 | 18 | 18 |
| V/V158 | 0 | 1 | 2 | 2 | 2 | 2 | 3 | 3 | 3 |

**Probable AMR**

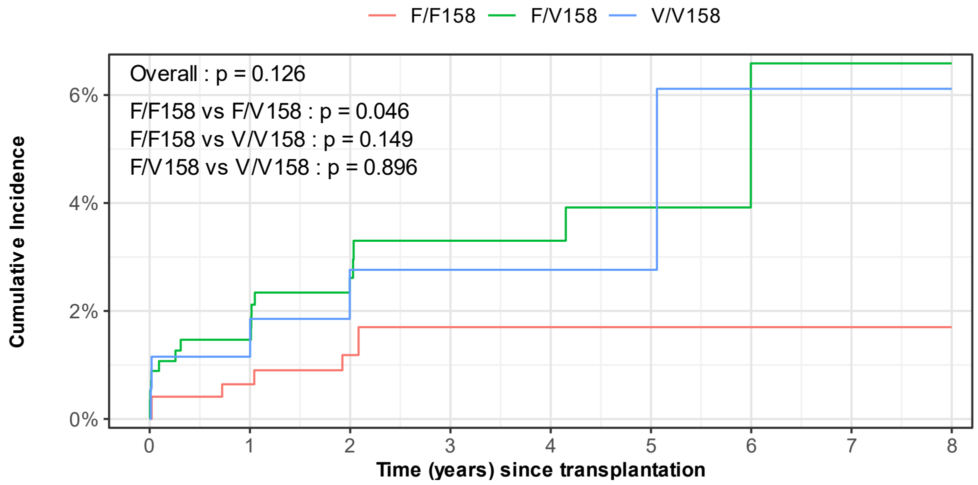


At Risk

| | 0 | 1 | 2 | 3 | 4 | 5 | 6 | 7 | 8 |
|---|---|---|---|---|---|---|---|---|---|
| F/F158 | 498 | 408 | 293 | 150 | 132 | 103 | 37 | 29 | 22 |
| F/V158 | 572 | 464 | 346 | 176 | 160 | 113 | 35 | 28 | 20 |
| V/V158 | 178 | 141 | 101 | 53 | 45 | 38 | 14 | 9 | 8 |

Events

| | 0 | 1 | 2 | 3 | 4 | 5 | 6 | 7 | 8 |
|---|---|---|---|---|---|---|---|---|---|
| F/F158 | 0 | 3 | 5 | 6 | 6 | 6 | 6 | 6 | 6 |
| F/V158 | 0 | 8 | 13 | 15 | 15 | 16 | 17 | 17 | 17 |
| V/V158 | 0 | 2 | 4 | 4 | 4 | 4 | 5 | 5 | 5 |

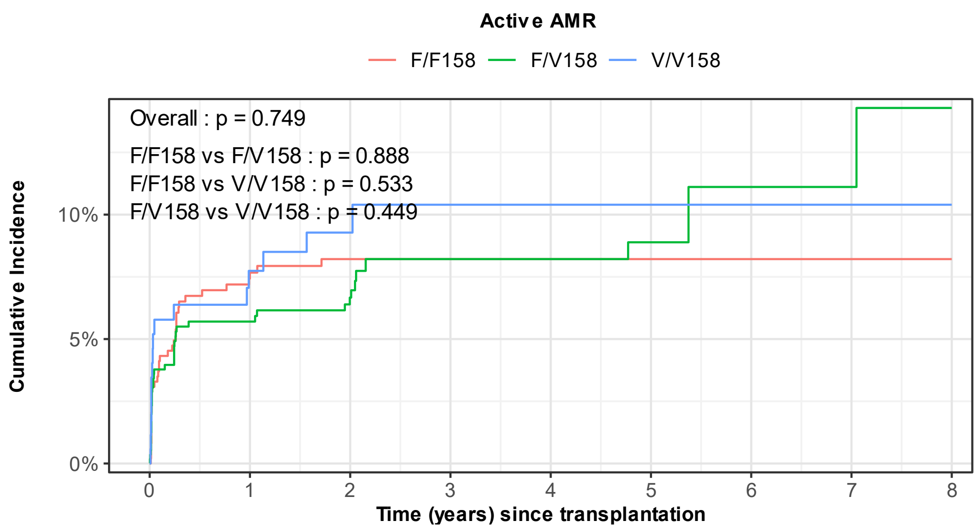


At Risk

| | | | | | | | | | |
|---|---|---|---|---|---|---|---|---|---|
| F/F158 | 498 | 383 | 276 | 141 | 124 | 96 | 36 | 27 | 21 |
| F/V158 | 572 | 443 | 330 | 163 | 147 | 105 | 35 | 28 | 20 |
| V/V158 | 178 | 133 | 94 | 48 | 41 | 34 | 13 | 8 | 7 |

Events

| | | | | | | | | | |
|---|---|---|---|---|---|---|---|---|---|
| F/F158 | 0 | 36 | 38 | 38 | 38 | 38 | 38 | 38 | 38 |
| F/V158 | 0 | 31 | 35 | 39 | 39 | 40 | 41 | 41 | 42 |
| V/V158 | 0 | 13 | 15 | 16 | 16 | 16 | 16 | 16 | 16 |

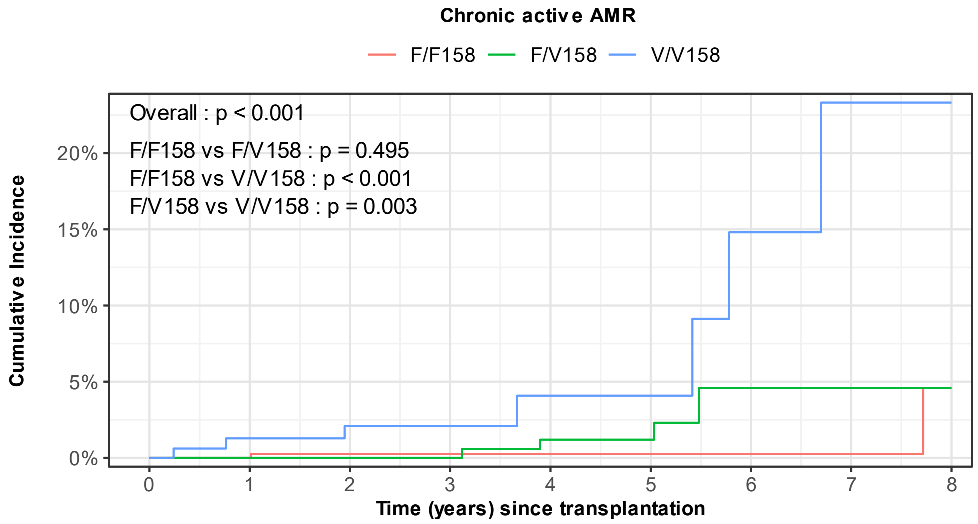


At Risk

| | | | | | | | | | |
|---|---|---|---|---|---|---|---|---|---|
| F/F158 | 498 | 410 | 294 | 150 | 132 | 103 | 37 | 28 | 21 |
| F/V158 | 572 | 470 | 354 | 180 | 161 | 115 | 36 | 28 | 20 |
| V/V158 | 178 | 142 | 101 | 54 | 46 | 39 | 14 | 9 | 8 |

Events

| | | | | | | | | | |
|---|---|---|---|---|---|---|---|---|---|
| F/F158 | 0 | 0 | 1 | 1 | 1 | 1 | 1 | 1 | 2 |
| F/V158 | 0 | 0 | 0 | 0 | 2 | 2 | 4 | 4 | 4 |
| V/V158 | 0 | 2 | 3 | 3 | 4 | 4 | 6 | 7 | 7 |

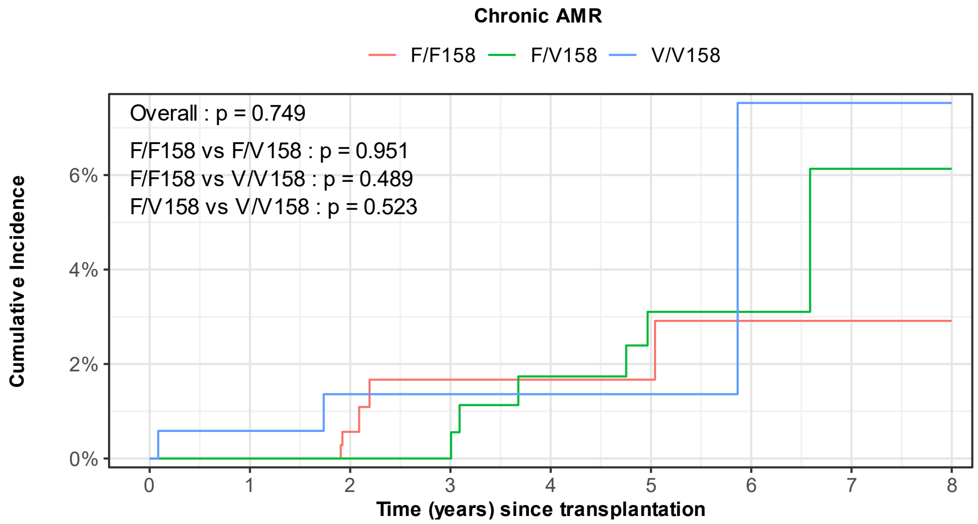


At Risk

| | 0 | 1 | 2 | 3 | 4 | 5 | 6 | 7 | 8 |
|---|---|---|---|---|---|---|---|---|---|
| F/F158 | 498 | 410 | 294 | 149 | 132 | 103 | 37 | 28 | 21 |
| F/V158 | 572 | 470 | 354 | 180 | 161 | 115 | 36 | 28 | 20 |
| V/V158 | 178 | 142 | 101 | 54 | 46 | 39 | 15 | 10 | 8 |

Events

| | 0 | 1 | 2 | 3 | 4 | 5 | 6 | 7 | 8 |
|---|---|---|---|---|---|---|---|---|---|
| F/F158 | 0 | 0 | 2 | 4 | 4 | 4 | 5 | 5 | 5 |
| F/V158 | 0 | 0 | 0 | 0 | 3 | 5 | 5 | 6 | 6 |
| V/V158 | 0 | 1 | 2 | 2 | 2 | 2 | 3 | 3 | 3 |

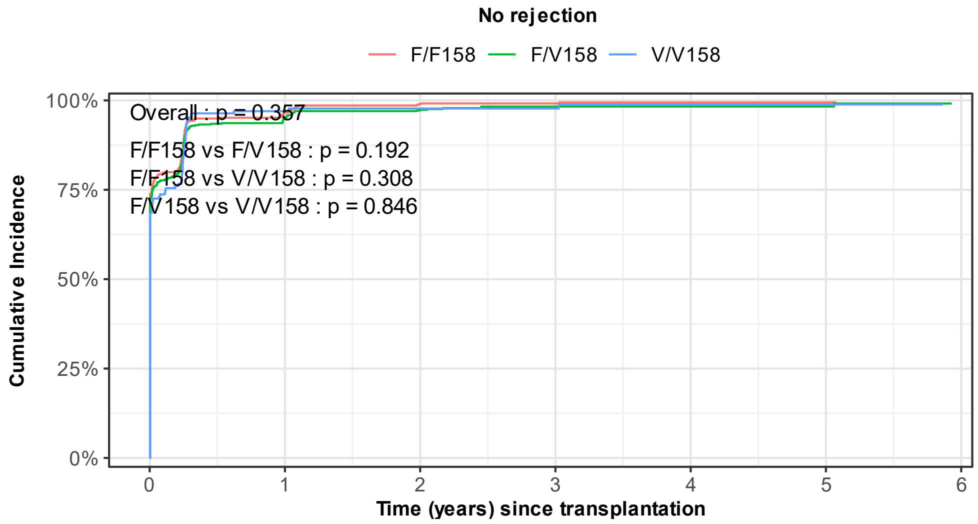


At Risk

| | 0 | 1 | 2 | 3 | 4 | 5 | 6 |
|---|---|---|---|---|---|---|---|
| F/F158 | 498 | 17 | 4 | 3 | 2 | 1 | 0 |
| F/V158 | 572 | 26 | 15 | 4 | 4 | 2 | 0 |
| V/V158 | 178 | 5 | 3 | 2 | 1 | 1 | 0 |

Events

| | 0 | 1 | 2 | 3 | 4 | 5 | 6 |
|---|---|---|---|---|---|---|---|
| F/F158 | 0 | 480 | 491 | 492 | 493 | 493 | [illegible] |
| F/V158 | 0 | 542 | 553 | 557 | 557 | 557 | [illegible] |
| V/V158 | 0 | 170 | 171 | 171 | 172 | 172 | [illegible] |

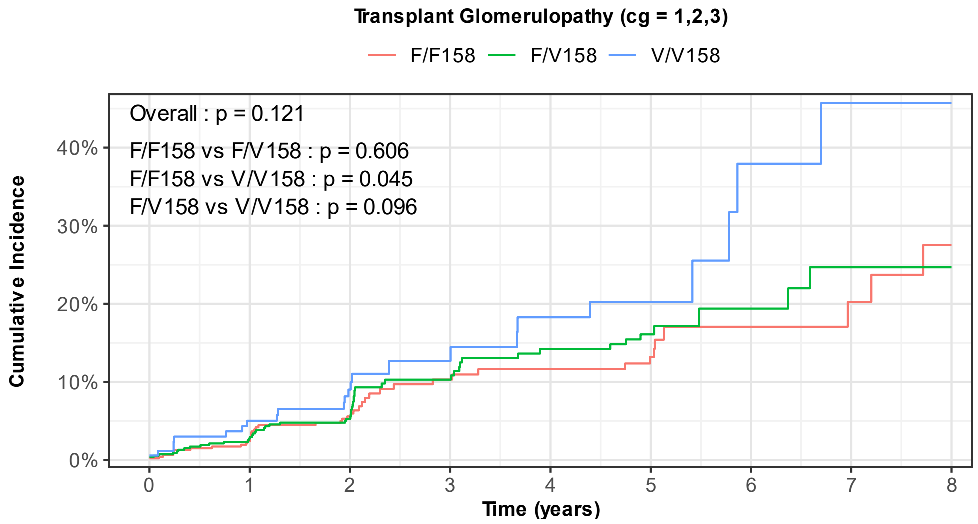


At Risk

| | 0 | 1 | 2 | 3 | 4 | 5 | 6 | 7 | 8 |
|---|---|---|---|---|---|---|---|---|---|
| F/F158 | 498 | 401 | 282 | 142 | 124 | 97 | 33 | 25 | 19 |
| F/V158 | 572 | 458 | 335 | 167 | 147 | 104 | 32 | 25 | 18 |
| V/V158 | 178 | 136 | 95 | 49 | 42 | 35 | 10 | 7 | 7 |

Events

| | 0 | 1 | 2 | 3 | 4 | 5 | 6 | 7 | 8 |
|---|---|---|---|---|---|---|---|---|---|
| F/F158 | 1 | 11 | 23 | 32 | 34 | 36 | 39 | 40 | 42 |
| F/V158 | 2 | 14 | 25 | 39 | 46 | 49 | 51 | 53 | 53 |
| V/V158 | 1 | 8 | 13 | 16 | 19 | 20 | 23 | 24 | 24 |

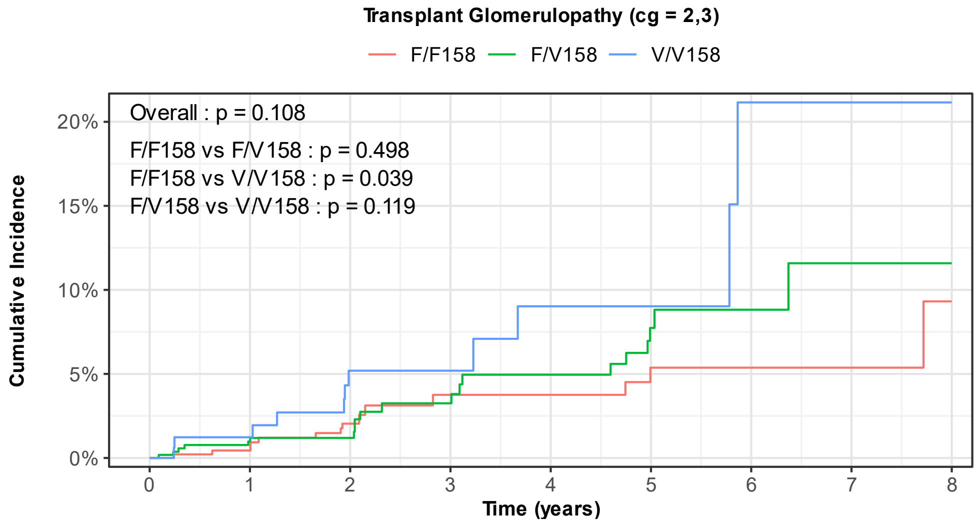


At Risk

| | 0 | 1 | 2 | 3 | 4 | 5 | 6 | 7 | 8 |
|---|---|---|---|---|---|---|---|---|---|
| F/F158 | 498 | 408 | 291 | 147 | 130 | 103 | 38 | 29 | 22 |
| F/V158 | 572 | 466 | 349 | 175 | 156 | 110 | 35 | 27 | 19 |
| V/V158 | 178 | 141 | 98 | 52 | 45 | 38 | 13 | 9 | 8 |

Events

| | 0 | 1 | 2 | 3 | 4 | 5 | 6 | 7 | 8 |
|---|---|---|---|---|---|---|---|---|---|
| F/F158 | 0 | 2 | 8 | 11 | 11 | 13 | 13 | 13 | 14 |
| F/V158 | 0 | 5 | 6 | 11 | 14 | 18 | 19 | 20 | 20 |
| V/V158 | 0 | 2 | 7 | 7 | 9 | 9 | 11 | 11 | 11 |

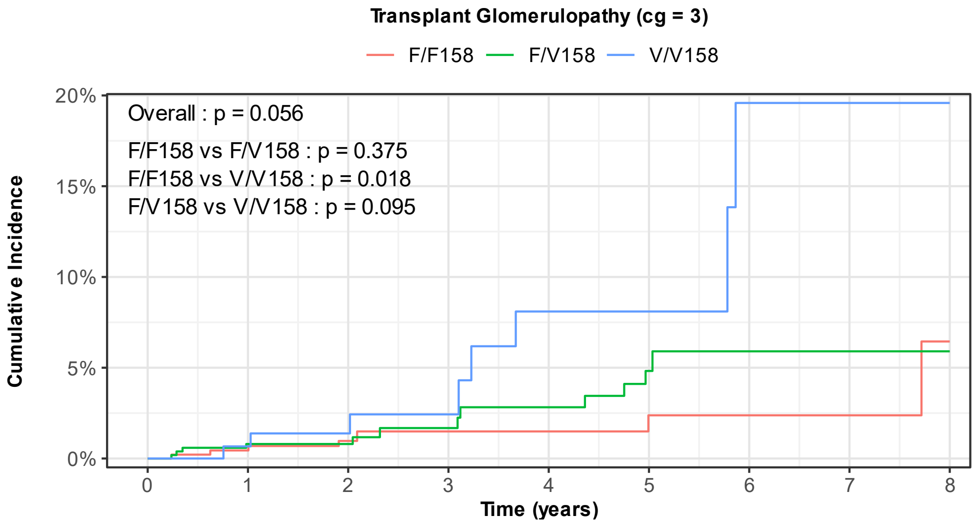


At Risk

| | 0 | 1 | 2 | 3 | 4 | 5 | 6 | 7 | 8 |
|---|---|---|---|---|---|---|---|---|---|
| F/F158 | 498 | 408 | 294 | 149 | 131 | 103 | 38 | 29 | 22 |
| F/V158 | 572 | 466 | 350 | 178 | 160 | 113 | 35 | 27 | 19 |
| V/V158 | 178 | 142 | 101 | 54 | 46 | 39 | 14 | 9 | 8 |

Events

| | 0 | 1 | 2 | 3 | 4 | 5 | 6 | 7 | 8 |
|---|---|---|---|---|---|---|---|---|---|
| F/F158 | 0 | 2 | 4 | 5 | 5 | 6 | 6 | 6 | 7 |
| F/V158 | 0 | 4 | 4 | 6 | 8 | 11 | 12 | 12 | 12 |
| V/V158 | 0 | 1 | 2 | 3 | 6 | 6 | 8 | 8 | 8 |

**Figure S2. Cumulative incidence of C4d without rejection stratified by *FCGR3A* F158V genotype with (upper panel) and without (lower panel) ABO-incompatible grafts.**

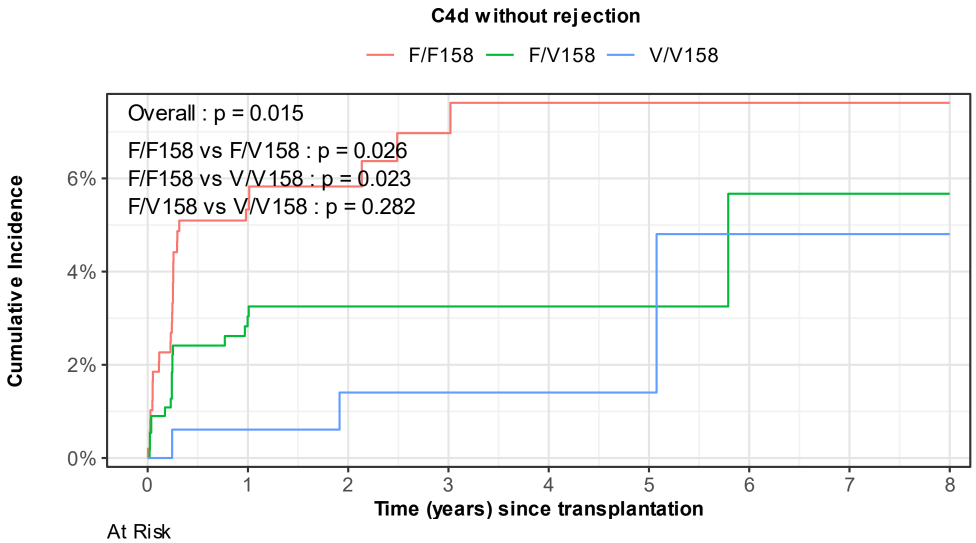


At Risk

| | 0 | 1 | 2 | 3 | 4 | 5 | 6 | 7 | 8 |
|---|---|---|---|---|---|---|---|---|---|
| F/F158 | 498 | 389 | 281 | 146 | 128 | 99 | 36 | 27 | 20 |
| F/V158 | 572 | 455 | 345 | 177 | 159 | 113 | 36 | 28 | 20 |
| V/V158 | 178 | 142 | 101 | 53 | 45 | 39 | 15 | 10 | 8 |

Events

| | 0 | 1 | 2 | 3 | 4 | 5 | 6 | 7 | 8 |
|---|---|---|---|---|---|---|---|---|---|
| F/F158 | 0 | 25 | 27 | 29 | 30 | 30 | 30 | 30 | 30 |
| F/V158 | 0 | 16 | 17 | 17 | 17 | 17 | 18 | 18 | 18 |
| V/V158 | 0 | 1 | 2 | 2 | 2 | 2 | 3 | 3 | 3 |

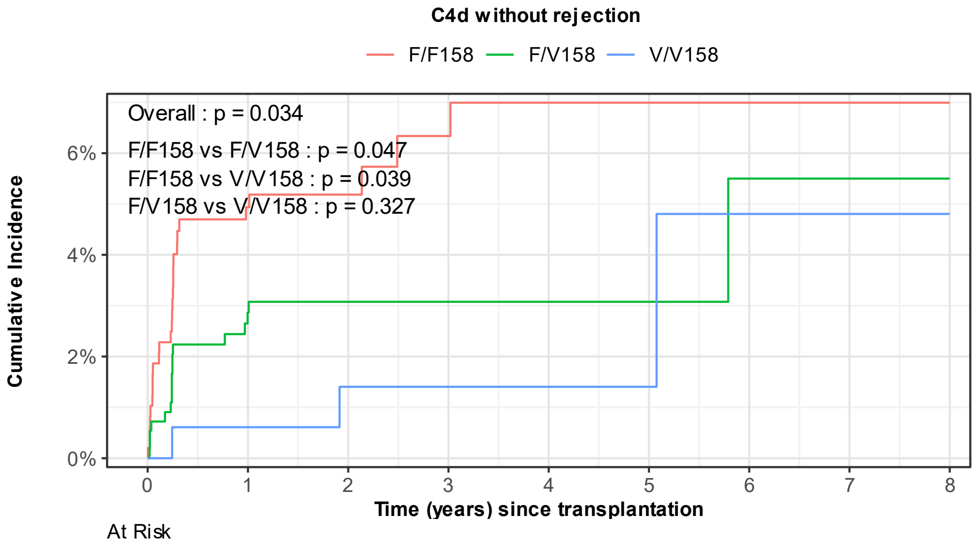


At Risk

| | 0 | 1 | 2 | 3 | 4 | 5 | 6 | 7 | 8 |
|---|---|---|---|---|---|---|---|---|---|
| F/F158 | 495 | 388 | 281 | 146 | 128 | 99 | 36 | 27 | 20 |
| F/V158 | 571 | 455 | 345 | 177 | 159 | 113 | 36 | 28 | 20 |
| V/V158 | 178 | 142 | 101 | 53 | 45 | 39 | 15 | 10 | 8 |

Events

| | 0 | 1 | 2 | 3 | 4 | 5 | 6 | 7 | 8 |
|---|---|---|---|---|---|---|---|---|---|
| F/F158 | 0 | 23 | 24 | 26 | 27 | 27 | 27 | 27 | 27 |
| F/V158 | 0 | 15 | 16 | 16 | 16 | 16 | 17 | 17 | 17 |
| V/V158 | 0 | 1 | 2 | 2 | 2 | 2 | 3 | 3 | 3 |

**Figure S3. Cumulative incidence of graft failure in the AMR spectrum cohort (C4d without rejection, probable AMR, active AMR, chronic active AMR and chronic AMR) stratified by *FCGR3A* F158V genotype.**

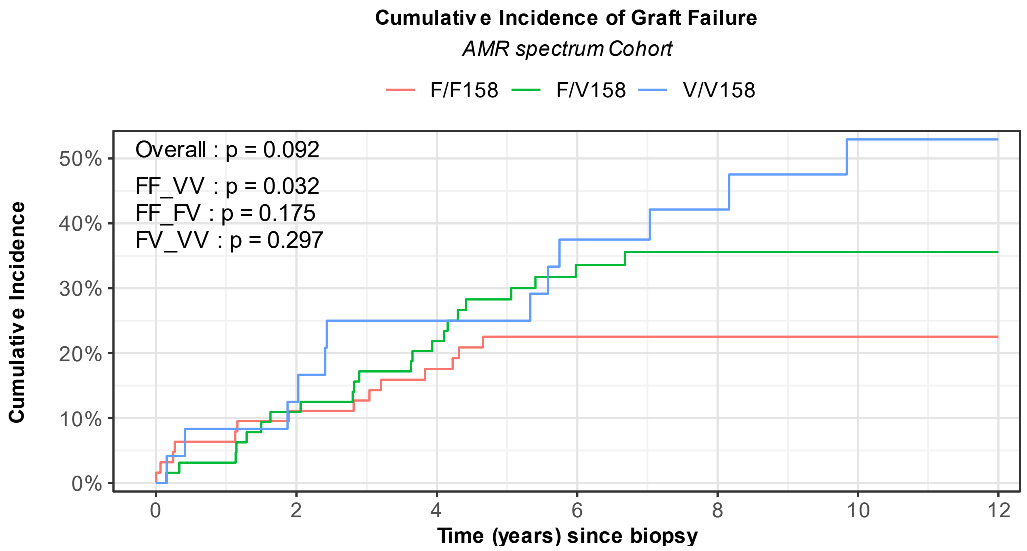


| | 0 | 2 | 4 | 6 | 8 | 10 | 12 |
|---|---|---|---|---|---|---|---|
| **At Risk** | | | | | | | |
| F/F158 | 63 | 51 | 44 | 33 | 18 | 12 | 10 |
| F/V158 | 64 | 55 | 45 | 29 | 22 | 16 | 8 |
| V/V158 | 24 | 18 | 13 | 9 | 6 | 2 | 2 |
| **Events** | | | | | | | |
| F/F158 | 0 | 7 | 11 | 14 | 14 | 14 | 14 |
| F/V158 | 0 | 7 | 14 | 21 | 22 | 22 | 22 |
| V/V158 | 0 | 3 | 6 | 9 | 10 | 12 | 12 |

**Figure S4. Conceptual figure depicting dissociation/association of complement activation and effective ADCC according to FCGR3A F158V genotype in antibody-mediated kidney transplant rejection.**

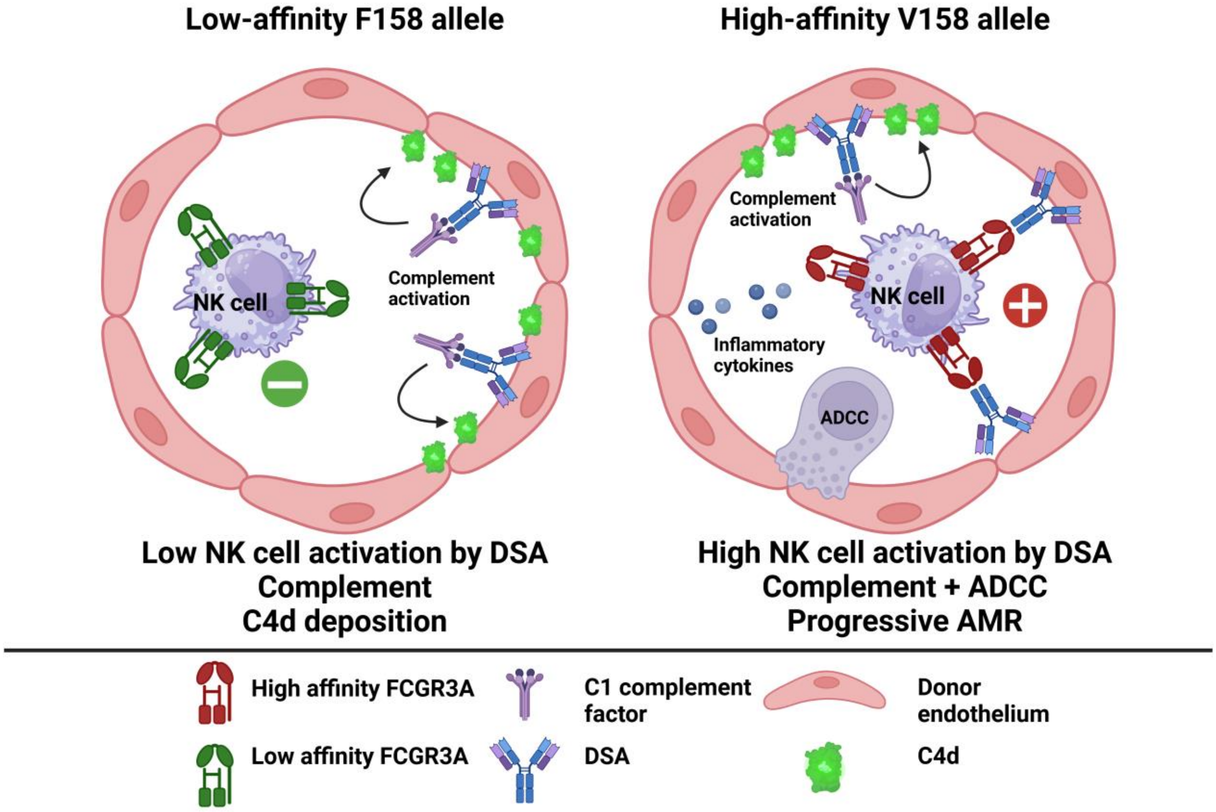
